# Contribution of endogenous and exogenous antibodies to clearance of SARS-CoV-2 during convalescent plasma therapy

**DOI:** 10.1101/2021.12.09.21267513

**Authors:** Maddalena Marconato, Irene A. Abela, Anthony Hauser, Magdalena Schwarzmüller, Rheliana Katzensteiner, Dominique L. Braun, Selina Epp, Annette Audigé, Jacqueline Weber, Peter Rusert, Emèry Schindler, Chloé Pasin, Emily West, Jürg Böni, Verena Kufner, Michael Huber, Maryam Zaheri, Stefan Schmutz, Beat M. Frey, Roger D. Kouyos, Huldrych F. Günthard, Markus G. Manz, Alexandra Trkola

## Abstract

Neutralizing antibodies are considered a key correlate of protection by current SARS-CoV-2 vaccines. The ability of antibody-based therapies, including convalescent plasma, to affect established disease remains to be elucidated. Only few monoclonal therapies and only when used at a very early stage of infection have shown efficacy. Here, we conducted a proof-of-principle study of convalescent plasma therapy in a phase I trial in 30 COVID-19 patients including immunocompromised individuals hospitalized early after onset of symptoms. A comprehensive longitudinal monitoring of the virologic, serologic, and disease status of recipients in conjunction with detailed post-hoc seroprofiling of transfused convalescent plasma, allowed deciphering of parameters on which plasma therapy efficacy depends. Plasma therapy was safe and had a significant effect on viral clearance depending on neutralizing and spike SARS-CoV-2 antibody levels in the supplied convalescent plasma. Endogenous immunity had strong effects on virus control. Lack of endogenous neutralizing activity at baseline was associated with a higher risk of systemic viremia. The onset of endogenous neutralization had a noticeable effect on viral clearance but, importantly, even after adjusting for their endogenous neutralization status recipients benefitted from therapy with high neutralizing antibody containing plasma.

In summary, our data demonstrate a clear impact of exogenous antibody therapy on the rapid clearance of viremia in the early stages of infection and provide directions for improved efficacy evaluation of current and future SARS-CoV-2 therapies beyond antibody-based interventions. Incorporating an assessment of the endogenous immune response and its dynamic interplay with viral production is critical for determining therapeutic effects.

**One Sentence Summary:** This study demonstrates the impact of exogenous antibody therapy by convalescent plasma containing high neutralizing titers on the rapid clearance of viremia in the early stages of SARS-CoV-2 infection.

## Introduction

The enormous burden that the SARS-CoV-2 pandemic poses to health and economic well-being can only be relieved if effective prevention and treatment options are available. While the development and introduction of effective SARS-CoV-2 vaccines has been successful at an unprecedented pace (1, 2), options for antiviral therapy to treat patients who develop severe coronavirus disease (COVID)-19 remain inadequate (3). Given the severe course of COVID-19 and the high mortality rate, especially among the elderly and those with concomitant diseases (4, 5), effective treatment options are urgently needed. Unfortunately, none of the newly developed or repurposed drugs, including remdesivir, has thus far demonstrated sufficient antiviral activity in advanced COVID-19 (3, 6).

Neutralizing antibodies are recognized as a principal correlate for protection induced by SARS-CoV-2 vaccines (7, 8) and have been considered for antiviral treatment as active component in convalescent plasma therapy (9-15) and as monoclonal antibody (mAb) therapeutics (16-18). Unless applied very early, in most clinical trials antibody-based SARS-CoV-2 therapies have not achieved the substantial disease-modulating effect initially hoped for. The influence of both plasma- and neutralizing mAb-based interventions appears to be limited to early infection at best (3, 6, 11, 19). Accordingly, therapeutic mAbs are thus far predominantly applied in non-hospitalized, early infection for persons at risk (20, 21).

Given the immediate availability of convalescent plasma and the urgent need for treatment, trials of convalescent plasma therapy were initiated shortly after the beginning of the pandemic, and lead to FDA approval of convalescent plasma therapy for early stage disease (22-26). However, despite numerous clinical trials, the utility of convalescent plasma in COVID-19 remains uncertain with most studies reporting no or only modest effects (23-25, 27-29), and few indicating a clear benefit (12, 13, 30-33). Meta-analyses, including randomized and non-randomized observational cohorts, suggest that convalescent plasma may be beneficial in a subset of patients (9-15). A randomized controlled trial reinforced a beneficial effect of convalescent plasma, particularly when high titers of neutralizing antibody are administered to elderly individuals (11). These findings are opposed by several other randomized controlled trials that failed to demonstrate the efficacy of convalescent plasma, particularly in patients who already have severe disease or require mechanical ventilation (24, 25, 27, 28, 34-36). The CONCOR-1 trial, a large multi-center randomized trial that randomized 938 patients, not only observed no benefit but identified a potentially harmful effect associated with non-neutralizing antibody activity in plasma (37).

The disparity of outcomes of convalescent plasma therapy trials remains difficult to comprehend. Individual studies differed greatly in terms of design, cohort size and measured outcome parameters. Differences in recipient disease stage and convalescent plasma characterization are limiting for direct outcome analyses and comparisons. As content of spike SARS-CoV-2 antibodies, especially neutralizing antibodies, have not been consistently monitored, it is almost impossible to decipher their effects and compare studies.

It is important to recognize that the activity of transfused antibodies is limited to a short period of time owing to the natural half-life of antibodies while mortality, the primary outcome parameter used across studies to assess efficacy of plasma therapy, is recorded at a considerable time-delay after transfusion. Therefore alternative outcome and analysis parameters need to be established in order to determine the principal efficacy of plasma antibody therapy and to ascertain the absence of detrimental effects. Since comprehensive assessment of clinical and laboratory parameters is not feasible in large-scale trials, small investigational proof-of-principle studies are needed to define suitable outcome and parameter measures. With this goal in mind, we used the framework of a phase I study to conduct a proof-of-principle study (NCT04869072) to evaluate the safety of convalescent plasma therapy and explore donor plasma antibody and outcome parameters for future efficacy studies. Novel to previous studies, the study population comprised 37% immunocompromised patients, a clearly underrepresented population in COVID clinical trials. Next to the conventional outcome and clinical parameters, we focused on a comprehensive assessment of the recipient seroconversion status and the SARS-CoV-2 antibody profile of donor plasma to elucidate which parameters are associated with viral clearance.

## Results

### Proof-of-principle study design

To investigate the potential of convalescent plasma therapy, we conducted a non-randomized, open-label, phase I clinical trial (NCT04869072) that included a comprehensive SARS-CoV-2 antibody profiling of donor plasma alongside a longitudinal monitoring of laboratory and clinical parameters (Fig.1 A, Table S1-S4). The aim of this proof-of-principle study was to ascertain safety of plasma therapy (primary outcome) and by extensive monitoring of parameters, to determine which effect transfused SARS-CoV-2 plasma antibodies have on the virological and disease status (secondary outcomes) (Table S4). Ranked as a first-in-human study by Swiss authorities, the safety focus was a requirement and excluded the formation of a no-treatment group. We accordingly tailored the study to allow a within-study efficacy assessment. Patients received three units of plasma (200ml each) of a single donor on three consecutive days followed by an extensive clinical and laboratory marker monitoring over 70 days (Fig. 1A). Consecutive administrations of smaller plasma volumes (200 ml each) were chosen to limit the risk of transfusion-related adverse events while allowing high dosing of convalescent plasma. The study protocol did not specify a threshold for SARS-CoV-2 serum antibodies in donor plasma for several reasons. At the time of study initiation, April 2020, the role of protective, neutralizing antibodies had not been ascertained and validated serology and neutralization tests were not yet available. Setting arbitrary thresholds for SARS-CoV-2 reactivity without knowing relevant protective levels was thus considered as problematic, as if wrongly set, this may limit the potential to retrieve information on the therapeutic effect of SARS-CoV-2 antibodies. The study design thus allowed for inclusion of plasma donors without prior screening for SARS-CoV-2 antibody levels. This ascertained that plasma used for therapy would capture a range of SARS-CoV-2 levels enabling a post-hoc analysis of the influence of antibody dose on outcome.

**Fig. 1:**
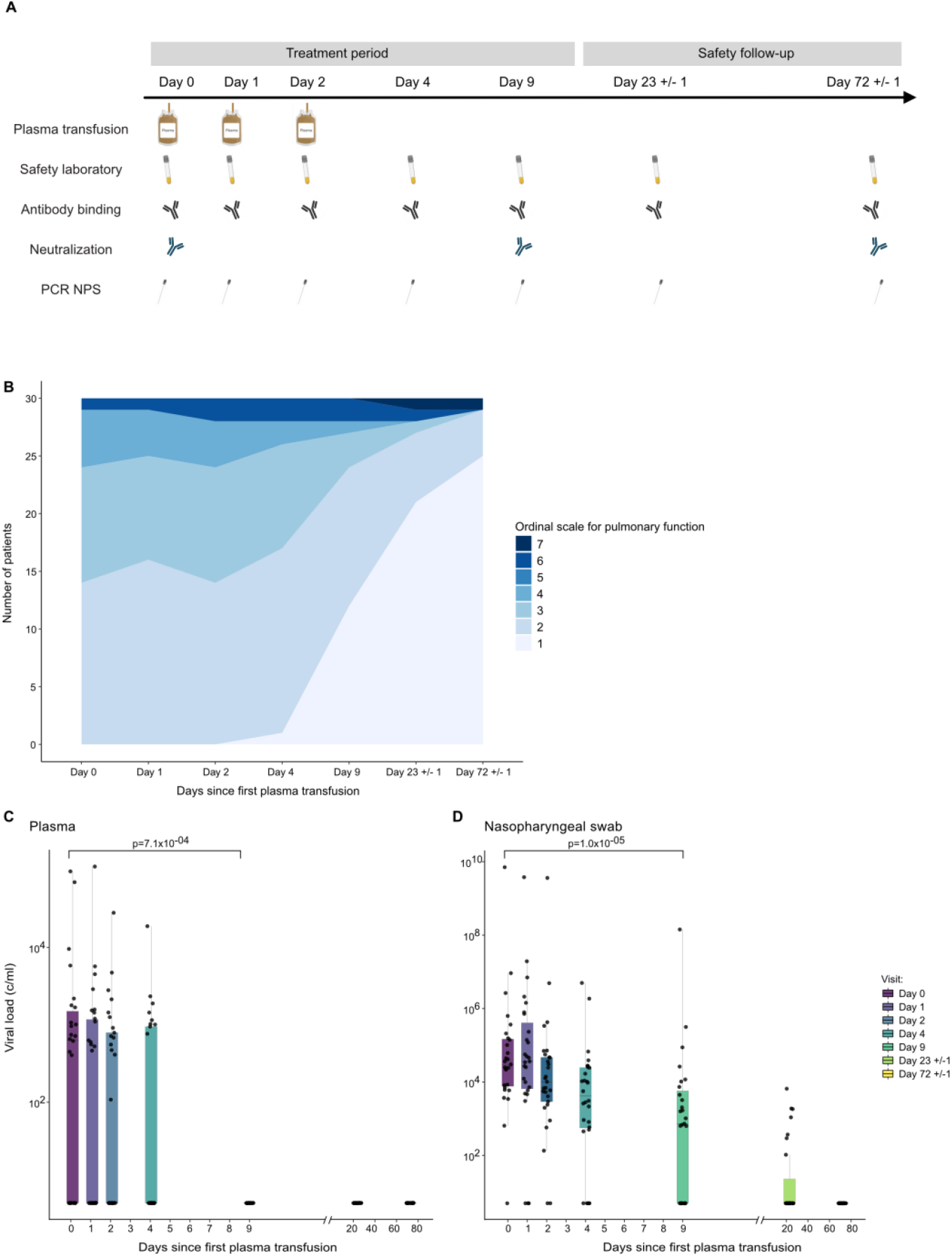
Study design, clinical and virological assessment. (A) Schematic depiction of the study design including timeline of consecutive treatment with convalescent plasma units, clinical and laboratory assessments. PCR from nasopharyngeal swab (PCR NPS). Figure created with BioRender.com (B) Longitudinal clinical assessment of trial participants, n=30) with a seven-category ordinal scale for pulmonary function. 1: Usual activities with minimal/no symptoms. 2: No supplemental oxygen; symptomatic and unable to undertake usual activities. 3: Supplemental oxygen <4L/min. 4: Supplemental oxygen >=4L/min. 5: Non-invasive ventilation or high-flow oxygen. 6: Invasive ventilation, Extracorporeal membrane oxygenation (ECMO), mechanical circulatory support. 7: Death. (C-D) Assessment of viral load. Longitudinal viral RNA concentrations (copies/ml) in plasma (C) and nasopharyngeal swab (NPS) (D) in trial participants (n=30).

### Study population and baseline characteristics

Between April and December 2020 a total of 30 SARS-CoV-2-infected patients, hospitalized with COVID-19, were enrolled at a single trial center, the University Hospital Zurich, Switzerland according to the approved study inclusion and exclusion criteria (Table S2). All trial participants showed radiological signs of COVID-19 pneumonia at inclusion with 18/30 (60%) requiring oxygen supplementation but not treatment in an intensive care unit (ICU). Baseline characteristics at inclusion (Table 1, Table S5) were typical for COVID-19 and reflected the demographic distribution observed for hospitalized patients in Switzerland in 2020 (38, 39). Thirty-four percent of patients had immunosuppression, including immunodeficiency (17%), cancer (10%), and solid organ transplantation (7%). The median age at inclusion was 63.5 years (IQR, 58.2-68.5), 10/30 participants (33%) were women, 22/30 participants (73%) presented with one or more comorbidities.

**Table 1:**
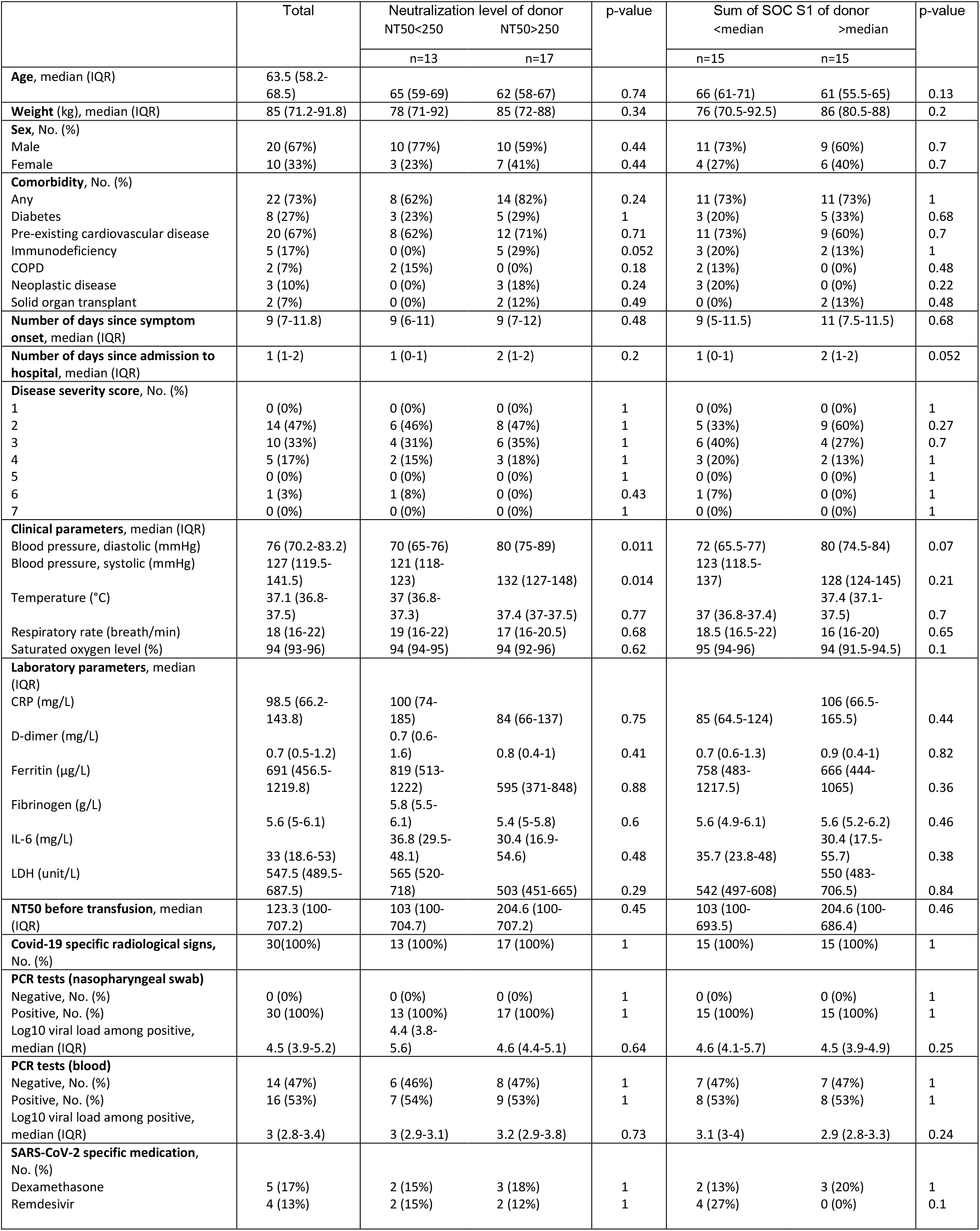
Baseline characteristics of trial participants (plasma recipients)

During the trial period, mostly SARS-CoV-2 lineages derived from B.1, harboring D614G but otherwise closely related to the original Wuhan-1 HU-1 strain (MN908947.3), were prevalent in Switzerland (www.gisaid). Full genome sequencing of SARS-CoV-2 in NPS of 26 plasma recipients confirmed this (Fig. S2, Table S5).

In addition to plasma therapy, all patients received the standard COVID-19 treatment recommended in Switzerland at the respective time of admission. This was initially limited to symptomatic control, supportive care and oxygen therapy and was later extended to include therapy with remdesivir (3, 19, 40) and/or steroids (41) when these options became available in Switzerland (Table 1, Table S5). Five patients (17%) received systemic steroid therapy, four (13%) were treated with remdesivir, and one (3.3%) received combination therapy with systemic steroids and remdesivir (Table 1, Table S5).

### Convalescent plasma therapy and safety assessment

Participants received three 200ml plasma donations of the same AB0-compatible plasma donor on three consecutive days. Transfusions were supervised by expert hematologists. No transfusion-related adverse events occurred within the first 72 hour post administration and during follow-up visits (Table S6). Adverse events, that were ranked as related to COVID-19 or other underlying diseases but not plasma therapy, were observed in 6/30 individuals (20%) which is in the range expected for the included COVID-19 stage (Table S7 (26)).

The key outcome measured across current SARS-CoV-2 convalescent plasma trials is mortality, assessed 3-4 weeks post transfusion (23, 24, 42). In our trial, one patient (Pat 15, Table S5) suffering from chronic lymphocytic leukemia died from bacterial, hospital acquired pneumonia by day 12. No other deaths occurred by study completion, resulting in an overall mortality rate of 3.3% (Table S7) within 72 days after study enrolment.

While decreasing mortality in COVID-19 is the ultimate goal, we sought to further include outcome measures that allow a gradual assessment of disease progression and cure. To this end we longitudinally assessed patient’s health status by a seven-category ordinal scale for pulmonary function as described (43, 44). The function score improved gradually, with 25/30 (83%) patients reaching full pulmonary function by study completion (Fig. 1B, Table S8). Median duration of hospitalization was 8 days (IQR: 6-13). Only two participants required intensive care over the trial period and needed mechanical ventilation (Fig. 1B, Table S5 and S7). Notably, we observed overall a rapid improvement in respiratory rate, oxygen saturation and body temperature at day 9 since first convalescent plasma unit, i.e. one week after the last plasma dose (Fig. S1A, Table S9). Laboratory markers of inflammation progressively improved and were within the reference values at study completion (day 72, 10 weeks post transfusion) for the majority of participants (Fig. S1B). C-reactive protein (CRP) levels in particular showed a rapid decline following plasma therapy (Fig. S1B). Coagulopathy, indicated by increased D-dimer and fibrinogen levels, has been frequently observed in patients with COVID-19 and is associated with subsequent thromboembolic events and severe outcomes (45-49). Notably, fibrinogen was elevated at baseline in all patients but was already significantly decreased by day 4 (p= 0.0062, paired t-test), whereas D-Dimer levels were elevated only in a fraction of participants (21/28, 75%) and remained at comparable levels throughout.

To monitor virological improvement, we measured SARS-CoV-2 viral load in blood and nasopharyngeal swabs (NPS) (Fig. 1C-D). Median log10 baseline viral load in NPS was 4.5 (IQR: 3.9-5.2), with 16 individuals presenting with measurable SARS-CoV-2 viremia in plasma. Viral load in both specimens rapidly decreased in line with the normalization of clinical parameters (Fig. 1C-D).

### Antibody profiling of SARS-CoV-2 convalescent plasma

During the study, a total of 105 plasma donations were collected from convalescent male donors to ascertain the availability of AB0 compatible plasma (Fig. 2A, Table S10). Post-hoc analysis of the SARS-CoV-2 serological responses in this plasma cohort with the sensitive multiplex seroprofiling test ABCORA (50), showed a heterogeneous pattern prototypic for SARS-CoV-2 infection ranging from high responses with IgG, IgA and IgM reactivity to spike proteins (S1, RBD and S2) and nucleoprotein (N) to low reactivity (Fig. 2B). This pattern of high and low serum responses was confirmed by monitoring with the Elecsys Anti-SARS-CoV-2 S assay (Roche Diagnostics GmbH) that measures total immunoglobulin against RBD (Fig. 2C).

**Fig. 2:**
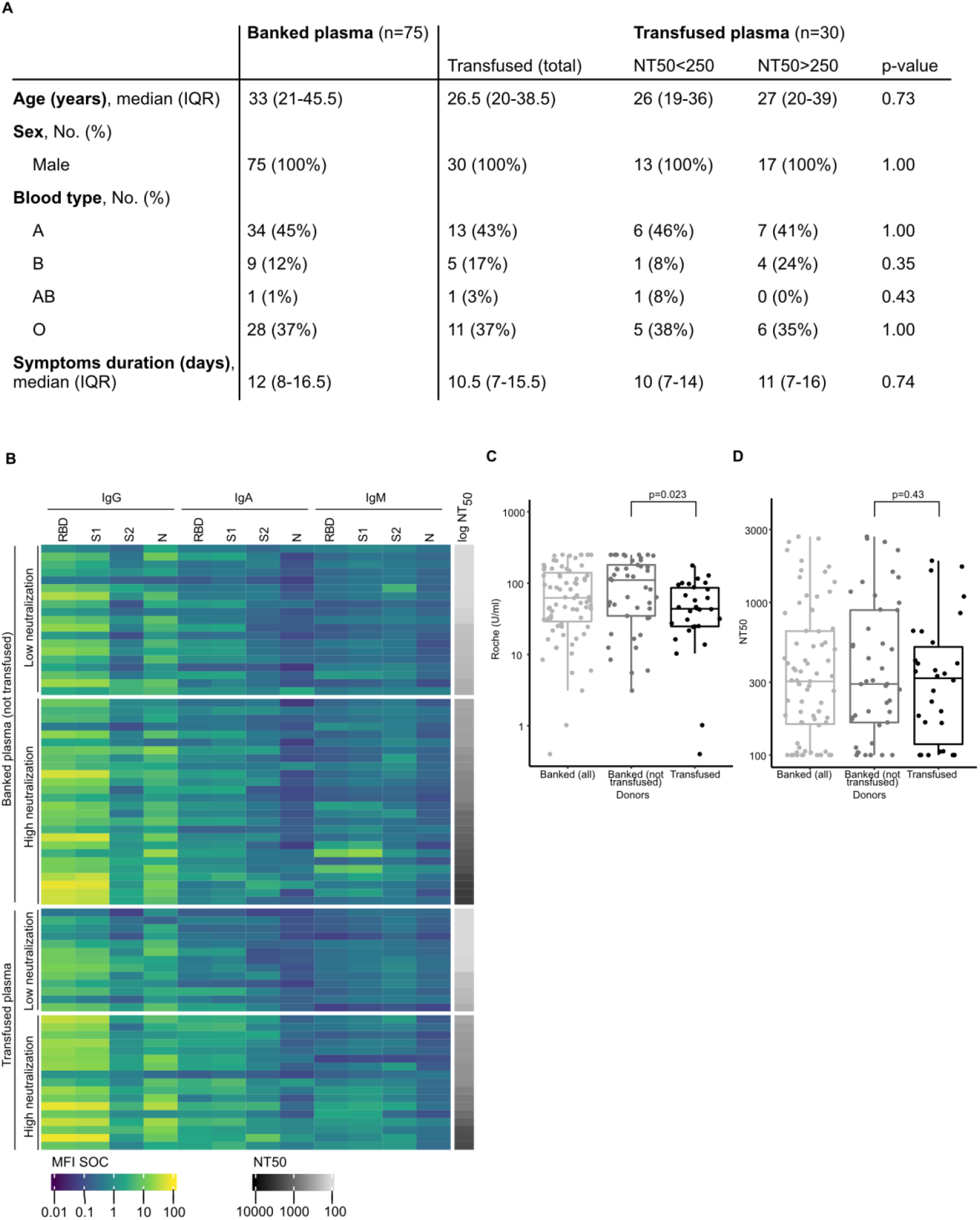
Characteristics of convalescent plasma donors. (A) Basic demographics of banked (n=75) and transfused (n=30) plasma from convalescent donors. Transfused plasma is further stratified in plasma containing low (NT50<250) and high (NT50>250) neutralizing activity. (B) Heatmap representing the seroreactivity to SARS-CoV-2 antigens in the SARS-CoV-2 ABCORA 2 test and 50% neutralization titers (NT50) titers against Wuhan-Hu-1 pseudotype post SARS-CoV-2 of all banked (n=75) and transfused (n=30) convalescent plasma. ABCORA binding reactivity is depicted in MFI-SOC (signal over cut off) values. Donor plasmas are stratified into high (NT50>250) and low (NT50<250) neutralization potency. (C) Antibody binding titers as assessed by Elecsys Anti-SARS-CoV-2 (S) assay (U/ml) of all, banked but not transfused, and transfused convalescent plasma donors. Unpaired t-test comparing the two groups shows significant binding antibody titer difference. (D) NT50 titers against Wuhan-Hu-1 pseudotype of all, banked but not transfused, and transfused convalescent plasma donors. Unpaired t-test comparing the two groups shows no significant difference (p=0.27).

Neutralization levels were examined against Wuhan-1 HU-1 as a measure of antiviral activity in donor plasma. Neutralizing titers in donor plasma ranged from no neutralization activity to a titer of 2700 (Fig. 2D).

### Impact of convalescent plasma antibody on viral clearance

Release from hospital in our trial depended not only on the health status but also on de-isolation rules that were adapted over time by authorities. Time to hospital discharge could therefore not be used as an endpoint. We therefore directly assessed the impact on virological improvement and investigated if and which SARS-CoV-2-specific donor plasma antibody parameters are associated with viral clearance. Neutralizing antibodies are the presumed active antiviral component of convalescent plasma. Effective treatment should therefore result in decreasing viral load.

In conformity with analyses conducted by the Expanded Access Protocol (EAP) and the Food and Drug Association (FDA) (51, 52), we classified high and low neutralizing plasma by a 50% neutralizing titer (NT50) of 250 and stratified patients accordingly. Baseline characteristics of trial participants receiving high-and low-titer plasma were found to be very comparable (Table 1). Treatment with highly neutralizing plasma was associated with faster viral clearance, as shown by a basic Kaplan-Meier analysis (p = 0.034) (Fig. 3A) but not with time to hospital discharge (Fig. S3). We next verified this result in a parametric model that allowed for interval censored data for the SARS-CoV-2 antibody measurement and adjusted for two recipient baseline parameters. We adjusted for viral load (measured in NPS) because higher viral loads are likely to require longer to clear. We also considered that viral clearance was mediated by both, the patient’s endogenous immune response and the supplied convalescent plasma. We also adjusted for comorbidities, as several trial participants had underlying diseases that can lead to impaired immune function (Table S5). The parametric model including viral load and comorbidity confirmed the effect of convalescent plasma content on viral clearance (adjusted hazard ratio (HR) = 3 [95% confidence interval (CI) 1.1-8.1], p = 0.026, Fig. 3BC). Three individuals were incapable of mounting an antibody response to SARS-CoV-2. Excluding these patients in a sensitivity analysis did not alter the result (adjusted hazard ratio (HR) = 2.71 [95% confidence interval (CI) 1.0-7.7], p = 0.046, Fig. S4AB). Four patients received in addition to convalescent plasma also the antiviral remdesivir, which may equally impact on viral clearance. Excluding remdesivir treated individuals from the hazard ratio analysis in a further sensitivity verification, we observed an even higher impact of plasma neutralizing activity on viral clearance (adjusted hazard ratio (HR) = 4.8 [95% confidence interval (CI) 1.6-14], p = 0.0056 Fig. S4CD). Equally, excluding both, remdesivir treated individuals and those incapable of mounting an antibody response (n=6) does not alter the outcome of the analysis (Fig. S4EF). Based on these analyses, we concluded that high neutralizing activity in convalescent plasma promotes rapid viral clearance.

**Fig. 3:**
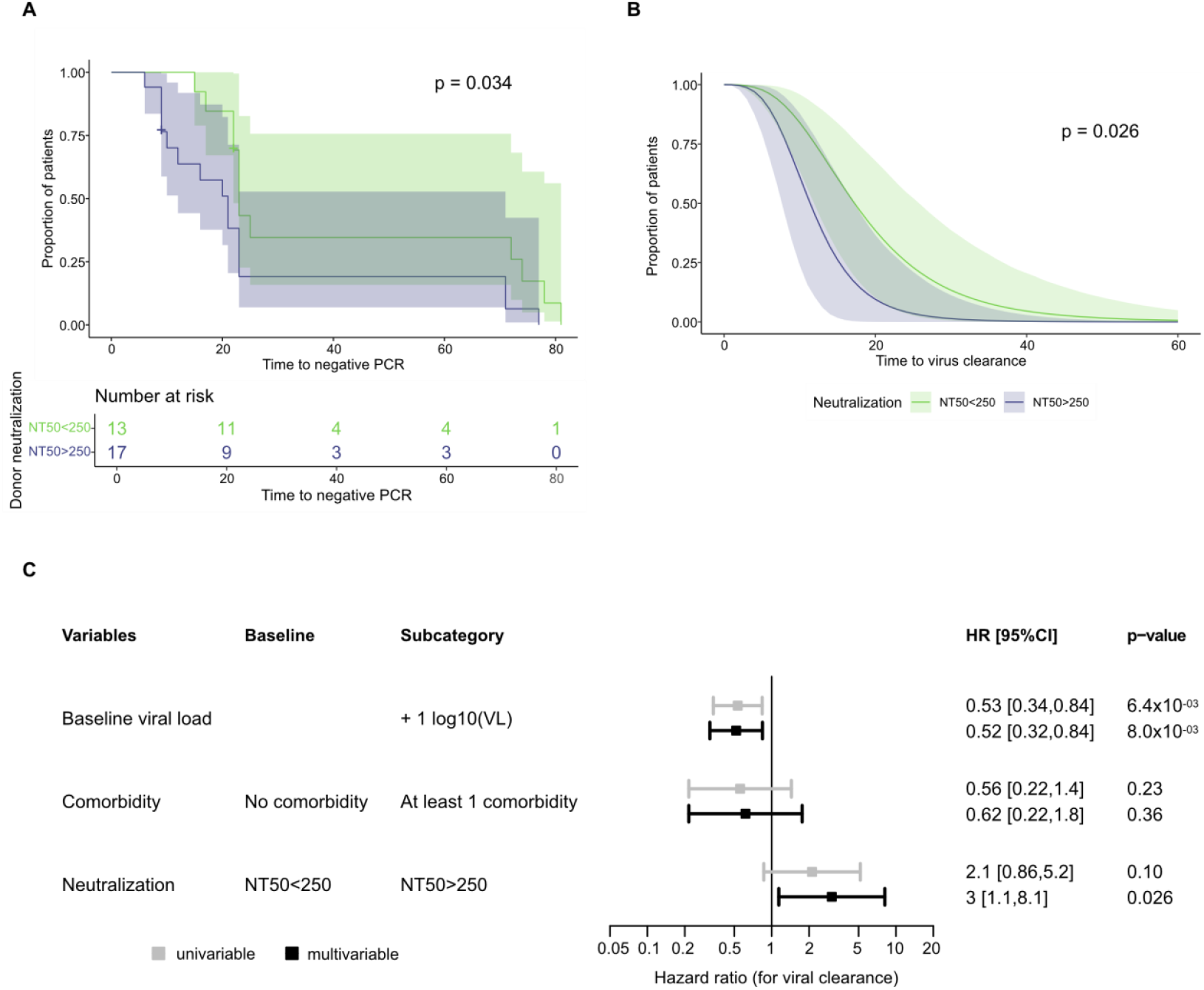
Treatment with highly neutralizing plasma leads to faster viral clearance. (A) Kaplan-Meier analysis and (B) survival function estimated with parametric model for interval-censored data assessing the time (in days) to viral clearance in trial participants (n=30) according to the level of neutralization potency in the transfused plasma. High neutralization activity NT50>250, low neutralization activity NT50< 250. (A) Kaplan Meier curves are compared by the log-rank test. (B) The parametric estimate is adjusted for the baseline NPS viral load and the presence of any comorbidity. The two survival curves for high and low neutralization trial participants correspond to the predicted viral clearance in individuals without comorbidity and with a baseline viral load equal to the median viral load observed among the 30 patients. (C) Forest plot corresponding to (B) showing the hazard ratios of the univariable (grey) and multivariable (black) model of time to viral clearance for neutralization level (low/high as in A and B), baseline viral load and the presence of comorbidity.

Considering that most convalescent plasma studies did not measure for neutralizing activity to assess antibody activity in convalescent plasma and instead relied on more readily available serologic assays, we next examined whether any of the antibody types among the 12 SARS-CoV-2 reactivities detected by ABCORA seroprofiling or the Elecsys S assay readout confirmed the finding for neutralizing antibodies. To this end, we performed a series of individual survival analyses assessing for each individual antibody reactivity and in addition composite reactivities the effect on time to viral clearance (summed IgG, IgA and IgM) (Fig. 4A, Fig. S5, Table S12). In each of these analyses, we stratified plasma according to the median reactivity into a high-and low-reactivity treatment group and again controlling for remdesivir treatment and immune suppression in sensitivity analyses. Most neutralizing SARS-CoV-2 antibodies target the RBD and the receptor-binding motif within the RBD, leaving only a comparatively small fraction of neutralization to S1 trimer-specific, spike N-terminal domain, and spike S2 antibodies (53-59). Accordingly, we initially focused on RBD responses but did not detect a differential effect of plasma on viral clearance when stratifying based on the Elecsys S assay that records binding to RBD (Fig. 4A, Fig. S6). This was in stark contrast to reactivities determined by the ABCORA seroprofiling test, where high S1 IgG, IgA, and IgM levels and high RBD IgA levels were associated with faster viral clearance irrespective of whether remdesivir treated individuals were included or not (Fig. 4A and Fig. S7). Particularly notable were IgA responses that displayed the highest impact on rapid viral clearance across all antigens. Due to their sequential evolution upon seroconversion, reactivities within an antibody class are expected to be correlated to a certain degree.

**Fig. 4:**
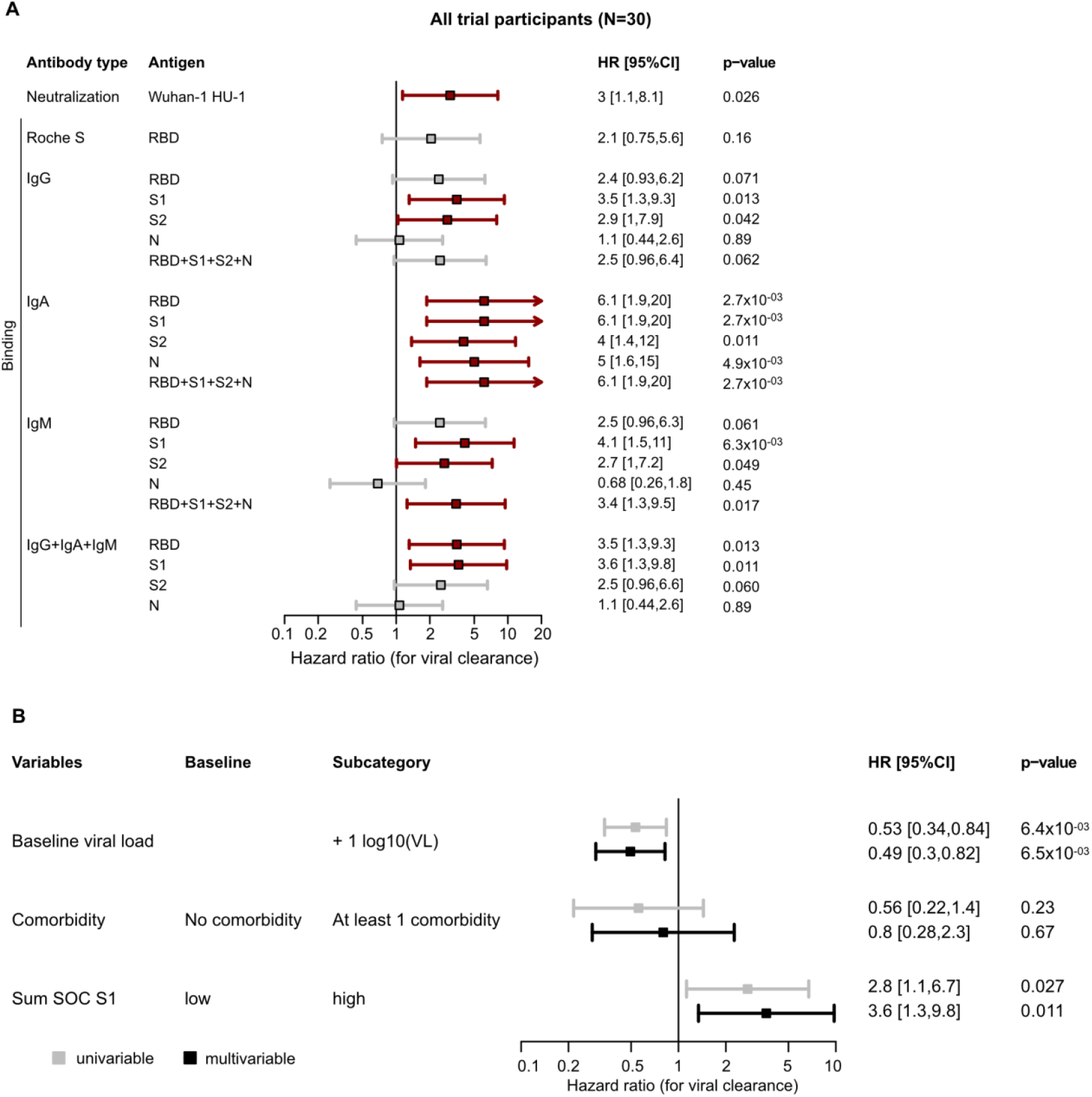
Spike specific binding and neutralizing antibodies are linked with rapid viral clearance. (A) Impact of antibody parameters on the time to viral clearance in multivariable parametric survival models including full trial cohort (n=30). Antibody reactivity of different antibody classes with SARS-CoV-2 antigens and neutralizing antibodies were included. Hazard ratios for individual antibody reactivities adjusted for the presence of comorbidity and the baseline viral load are shown. Significant results are marked in red. Low and high binding activity for each binding antibody parameter is stratified by the respective median binding reactivity. Low and high neutralization activity is stratified by a 50% neutralization titer of 250. (B) Forest plot depicting hazard ratios of univariable (grey) and multivariable (black) models of time to viral clearance including Elecsys Anti-SARS-CoV-2 (S) assay (U/ml). Multivariable analyses are corrected for baseline viral load and the presence of comorbidity. Low and high binding activity for each binding antibody parameter is stratified by the respective median binding reactivity.

This was also evident in our plasma donor cohort (Fig. S8). Based on the observed high association of S1 reactivities with viral clearance we selected the composite S1 value comprising IgG, IgA, and IgM S1 activity (sum S1), as parameters to be included in further analysis (Fig. 4B).

We further quantified the effect of plasma therapy on virus decay dynamics using censored regression models. We found that half-lives of virus load in NPS in recipients of high neutralizing plasma were shorter confirming the results from the hazard analysis (Fig. 5A) (half lifes: 1.5 days [95%CI: 1.1,2.2] when NT50>250 vs 1.0 days [95%CI: 0.69,2.0] when NT50<250, p=0.016). This effect was also evident when excluding remdesivir treated individuals and by stratifying for S1 reactivity (sum S1; Fig. 5B and Fig. S8).

**Fig. 5:**
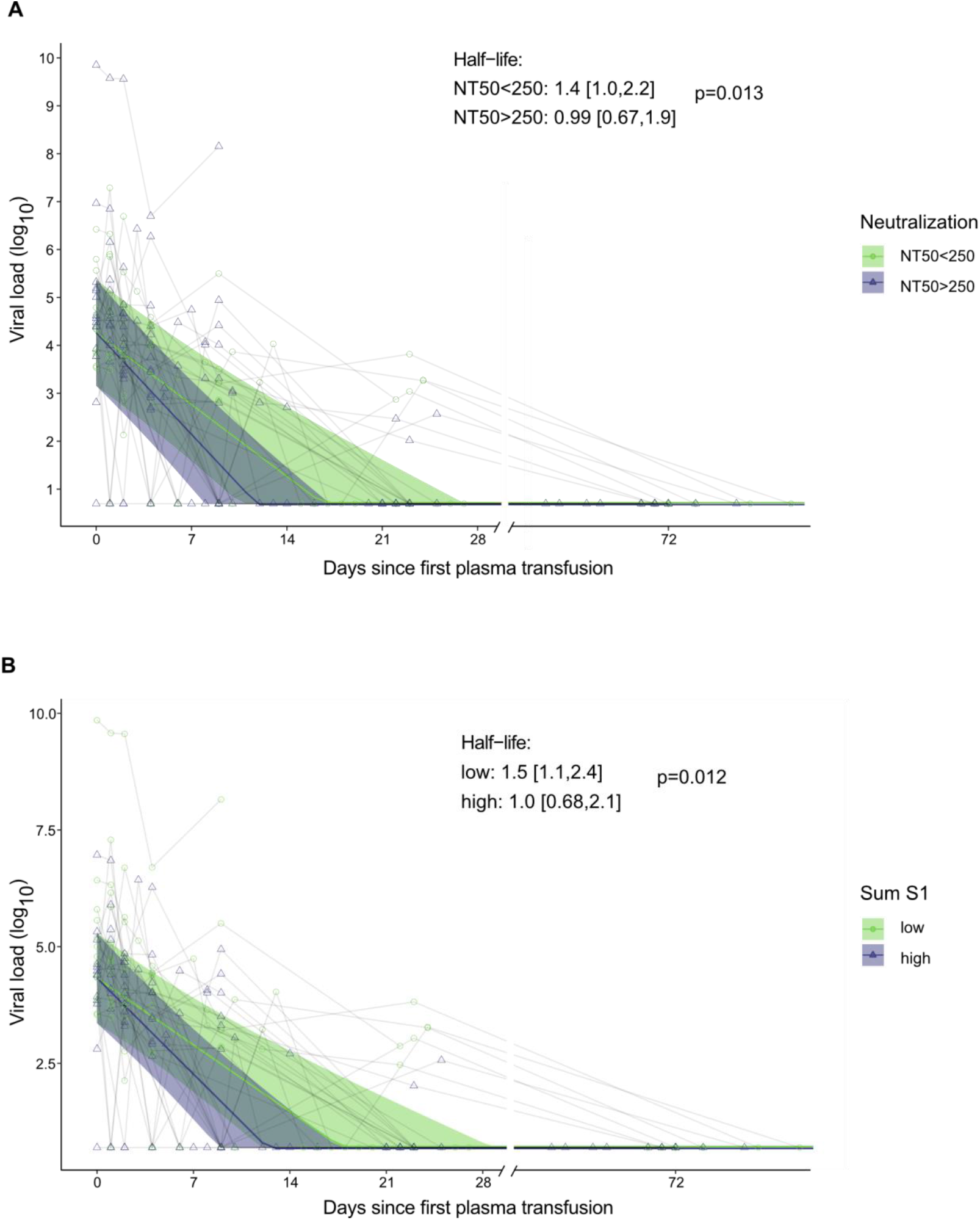
High neutralizing plasma leads to faster virus decay in nasopharyngeal swabs. (A and B) Censored regression model estimating decay rate of viral load (log10 viral load) of nasopharyngeal swabs in study participants (n=30) from time to treatment initiation according to (A) the level of neutralization (low neutralization NT50<250, light green; high neutralization, NT50>250, purple) or (B) level of binding defined by the ABCORA test sum of S1 SOC values. Low and high sum of S1 binding is stratified by the median binding reactivity. Significance was assessed using a two-sided t-test.

### Endogenous neutralizing antibodies control plasma viremia

Our trial focused on a comparatively early stage COVID-19 with severe pulmonary manifestation requiring hospitalization. The median interval between the onset of symptoms and the first transfusion was 9 days (IQR, 7-11.8 days) (Table 1). To rate the seroconversion status of trial participants, we monitored the evolution of SARS-CoV-2 antibodies at baseline and selected time points throughout the trial (Fig. 6AB, Fig. S10). Seroprofiling with the ABCORA test indicated a variable response at baseline ranging from low level partial responses to full blown seroconversion with high IgG anti-spike levels. Neutralization activity at baseline was likewise disparate with 12 individuals not reaching 50% neutralization activity (Fig. 6A). Overall, trial participants were relatively early in infection as illustrated by IgM responses at baseline (Fig. 6B). With the exception of three individuals with impaired immune function that induced no endogenous antibody response to SARS-CoV-2 (Fig. S11), neutralizing and binding antibodies increased in the majority of participants from baseline to day 9 and plateaued thereafter (Fig.6AB, Fig S10). Presence of neutralizing antibodies had an impact on the virological status of the recipient. Detectable SARS-CoV-2 in blood was inversely linked with neutralization activity and sum S1 levels at baseline (Fig. 6C, p= 0.0061), highlighting an effect of the endogenous neutralizing response on suppressing systemic viremia in the early phase of the infection. We therefore concluded that the endogenous serological status of the recipient, particularly neutralizing responses, and the virological status must be considered when evaluating SARS-CoV-2 antiviral therapies.

**Fig. 6:**
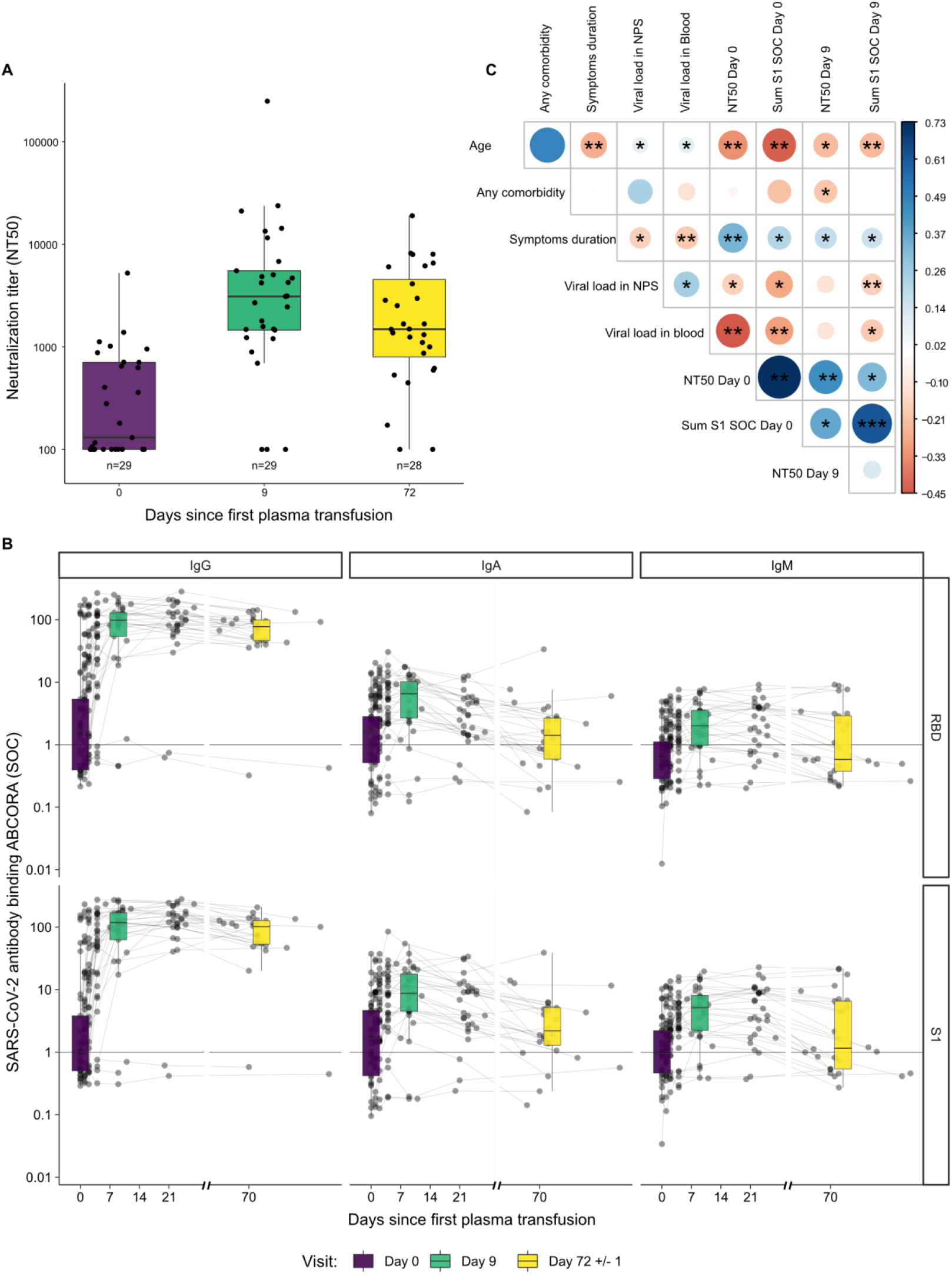
Endogenous neutralizing antibodies efficiently control plasma viremia. (A) Baseline endogenous 50% plasma neutralization titers (NT50) titers against Wuhan-Hu-1 pseudotyped virus was assessed longitudinally at day 0, day 9 and day 72. For each time point the number of patients with available sample are indicated. Box plots depict the interquartile ranges with vertical representing the minimum and maximum values. (B) Longitudinal binding antibody activity for study participants at baseline (day 0), day 9 and day 72 with the multiplex SARS-CoV-2 ABCORA 2.0 test. Sample numbers per time point as shown in (A). Depicted are signal over cutoff (SOC) values of IgG, IgA, IgM against RBD and S1. Box plots indicate the interquartile ranges with vertical lines representing the minimum and maximum values. (C) Spearman correlation matrix assessing correlation between age, comorbidities, viral load in NPS and blood, neutralization titer (NT50) and sum S1 SOC values at day 0 and day 9. Levels of significance are assessed by asymptotic t approximation of Spearman’s rank correlation. Color shading denotes correlation coefficient. Stars depict p<0.05.

### Impact of the endogenous serological status on the outcome of plasma therapy

Viral clearance in the context of the study needs to be viewed as the composite of the patients’ immunity and the activity of therapeutic plasma. We therefore investigated if endogenous and exogenous neutralizing antibody activity co-influences each other. For these analyses we excluded the three immune compromised individuals incapable of mounting antibody responses (Fig. S10). Neutralization titers significantly increased in both recipients of high and low neutralization plasma by day 9 since first convalescent plasma unit suggesting that by this time point the measured activity is dominated by the endogenous neutralization response and transfused antibodies have a marginal contribution (Fig. 7A-C). Titers in low NT plasma recipients significantly decreased (p=0.047) by day 70, whereas recipients of high NT plasma overall maintained endogenous activity (p=0.13) (Fig. 7A). Neutralization activity increased in immune competent individuals with and without endogenous neutralization activity at baseline over the observation period (Fig.7B). However, the increase in neutralization activity was significantly higher for individuals that already had neutralizing activity at baseline (p=0.038 at day 9) and these individuals maintained the gained activity longer (p=7.9×10^-3) (Fig. 7C).

**Fig. 7:**
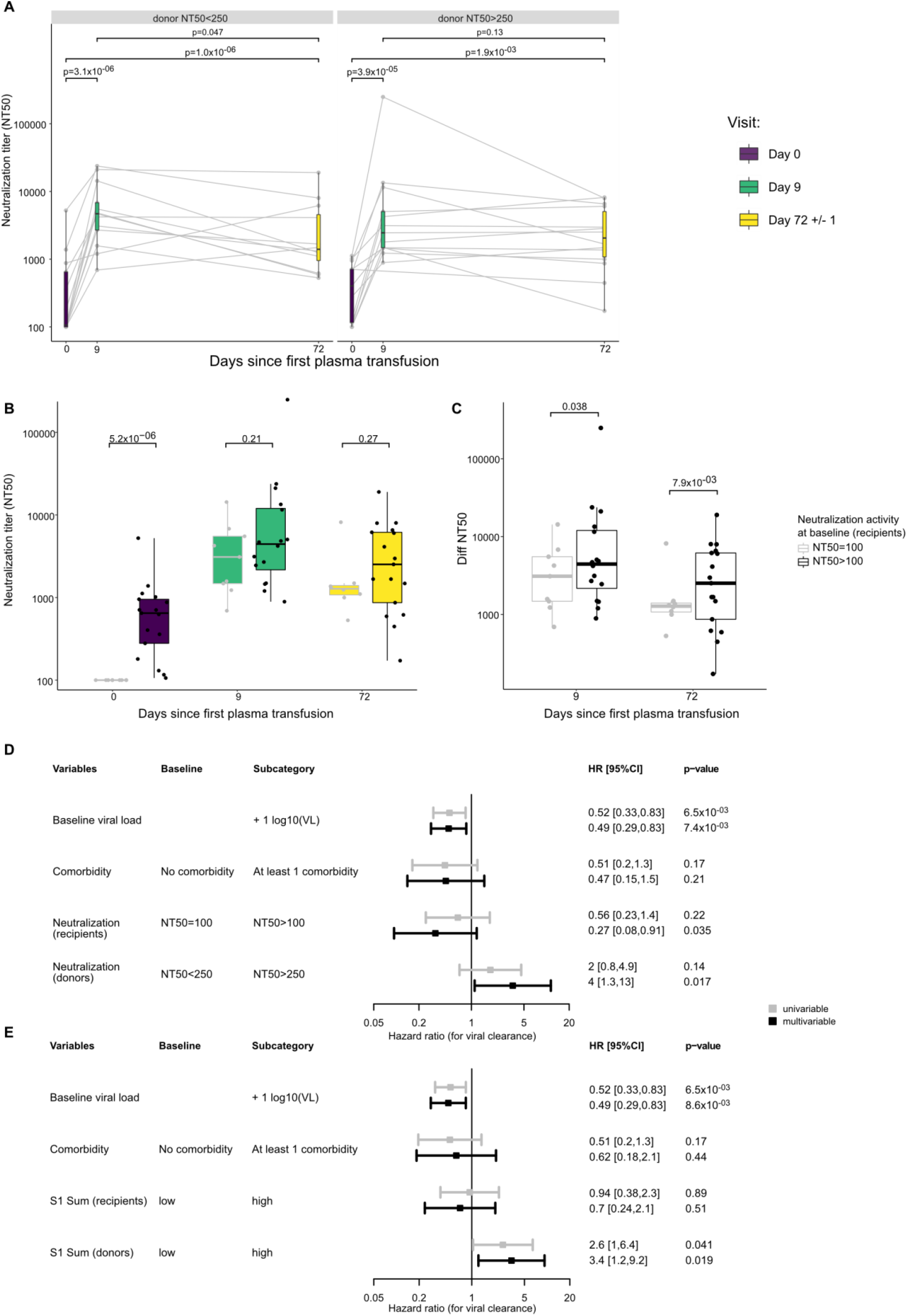
Influence of endogenous and convalescent plasma antibodies on viral clearance. (A) 50% Neutralization titers against Wuhan-Hu-1 pseudotype (NT50) at baseline (day 0, n= 29), day 9 (n= 29) and day 72 (n= 28) stratified by neutralizing levels of transfused convalescent plasma (left: low NT50, right: high NT50). Levels of significance are calculated by paired t-test. (B) NT50 post SARS-CoV-2 infection of plasma recipients at baseline (day 0, n = 29), day 9 (n = 29) and day 72 (n = 28) stratified by baseline neutralization activity; no neutralization (censored at NT50 = 100) or neutralization activity (NT50 >100). Levels of significance are assessed by unpaired t-test. (C) Increase of NT50 from baseline to Day 9 and Day 72, respectively. Trial participants are stratified by baseline neutralization activity; no neutralization (censored at NT50 = 100) or neutralization activity (NT50 >100). Levels of significance are assessed by unpaired t-test. (D,E) Forest plot showing hazard ratios of univariable and multivariable survival models of time to viral clearance. Both (D,E) test for baseline viral load and comorbidity; (D) tests additionally for donor and baseline endogenous plasma neutralization level; (E) tests additionally for donor and baseline endogenous plasma recipient S1 antibody level.

Finally, we tested for the combined effect of neutralization activity in the donor and recipient plasma on viral clearance. Specifically we adjusted the parametric survival model shown in figure 3C by accounting for recipient endogenous neutralization activity, in addition to baseline viral load, comorbidities and donor exogenous neutralization activity revealed effects of both convalescent plasma. High neutralization activity in convalescent plasma was associated with faster clearance, even after adjusting for recipients’ neutralization response at baseline (HR=4, [95% CI: 1.3, 13], p=0.017), demonstrating a positive impact of plasma therapy (Fig. 7D and Fig. S12). What may seem paradoxical at first glance, or even suggestive of an infection enhancing effect of the endogenous antibody response, is more plausibly explained by the rapid increase in neutralizing activity that we observed between baseline and day 9 of the study (Fig. 6A, Fig. 7BC). For patients that already have neutralizing antibodies at baseline, viral clearance is already ongoing and their virus loads at baseline need to be interpreted as declining. An increase in the endogenous neutralization response during the trial may thus not change their clearance rate substantially above levels at baseline. In contrast, for patients that mount an endogenous neutralization response only after trial entry, the measured viral clearance will be more notably impacted by the endogenous neutralization activity irrespective of the donor plasma they receive.

Evaluation of the effects of S1 antibody levels in both convalescent plasma and recipients at baseline corroborated the effects of convalescent plasma on viral clearance but not recipients’ S1 antibodies, thereby reaffirming the effects of endogenous neutralizing antibodies on viral clearance (Fig. 7E, Fig. S13).

## Discussion

In the present study we demonstrate the capacity of convalescent plasma therapy to induce rapid viral clearance. To date, most antibody-based therapies against COVID-19, particularly convalescent plasma therapies, have shown modest or no benefit (23-25). Highly diverse disease patterns in COVID-19 complicate the definition of a time window for treatment with neutralizing antibodies. The effect of passively administered antibodies is likely to be greatest at a stage when the patient’s own antibody response is not yet fully developed. Several trials with therapeutic SARS-CoV-2 antibody and convalescent plasma that showed efficacy, support this, suggesting that antiviral antibodies may be of most benefit when administered early in the course of infection (11, 12, 17, 42, 43, 60). While reducing mortality is the primary goal of any treatment, the fact that antibody-based therapeutics can be clinically impactful at early disease stages necessitates the measurement of additional outcomes. Although net effects of plasma treatment on mortality reduction could not be formally verified in the absence of a control group, the low mortality we observed in the trial is nonetheless noteworthy. The average mortality rates for hospitalized COVID-19 cases in the same time period in Switzerland of 9-13% were clearly higher (38, 39) allowing to exclude harmful effects of plasma treatment with respect to mortality in our study. Owing to its comparatively small size and specific design, the proof-of-principle study of convalescent plasma therapy for COVID-19 reported here, provided the opportunity to perform comprehensive longitudinal monitoring of recipients’ virologic, immunologic, and disease status. In conjunction with equally comprehensive post-hoc seroprofiling of donor plasma, this allowed to decipher parameters on which the efficacy of plasma therapy depends. Our data strongly argue for an influence of convalescent plasma on rapid viral clearance by neutralizing antibodies. The fact that S1 and RBD antibody activities of different antibody isotypes were associated with rapid viral clearance, supports a multifaceted action of antibodies beyond IgG driven neutralization, including antibody effector functions and neutralizing activity of IgA antibodies as previously suggested (61-63).

The results we present here provide key insights that may explain the apparent disparity of results of COVID-19 convalescent plasma trials. In particular, serologic tests may differ in their ability to detect relevant antibody activity, as demonstrated by the failure of the Elecsys S test to repeat findings confirmed by two other tests, ABCORA serology and neutralization measurements. This underscores the need for standardized assay systems and the definition of appropriate correlates and surrogate markers for antibody therapies including convalescent plasma. Serological profiling is a key element to define relevant parameters particularly in the context of the dynamic interplay of recipient and donor immunity as recently shown (64). In contrast to therapeutic antibodies, for which half-life can be optimized and which are administrable in high doses, the maximal activity of antibody therapy with convalescent plasma, which is commonly only given once, is inherently limited to a relatively short period after transfusion. Outcome measures, such as viral clearance are thus needed to enable the detection of treatment effects closer to transfusion. Rapid SARS-CoV-2 clearance must be considered as desirable, as therewith dissemination of the virus in the body will be halted and tissue damage limited. Yet, viral clearance may not always be linked with an immediate recovery. Depending on the progression of the disease and the state of inflamed organs and tissues, recovery periods may be extended. Antibody therapeutics may thus reliably record an impact on viral clearance but assessing the clinical impact of stopping virus replication will remain challenging as COVID-19 manifestation vary greatly.

The fact that the convalescent plasma in our study was provided during the build-up phase of the endogenous immunity may have been crucial in deciphering the effect of antibody therapy. Immune complexes formed by autologous antibodies and SARS-CoV-2, in particular anti-spike IgG have been implicated in causing severe lung immunopathology by inducing over-boarding inflammatory responses in alveolar macrophages (65-67). Of note, these effects have been linked to early spike IgG responses with aberrant glycosylation in individuals with severe COVID-19 (68). As glycosylation patterns normalize relatively rapidly (68), there is likely only a short window of time during seroconversion when immunopathologic spike IgG may circulate and cause severe disease. Seroconversion must thus be considered as a critical phase to target by therapeutics. In our trial, external, matured antibody was delivered at a time when the endogenous response was not yet fully complete. While immune complexes in early seroconversion may be detrimental if driven by aberrant glycosylated IgG, therapeutic antibodies and the fully matured antibody responses in convalescent plasma may substitute the endogenous response, opsonizing virus and destining it to clearance via Fc or complement receptor bearing phagocytes.

Antibody treatments need to be carefully evaluated for the absence of disease-exacerbating effects. Excessive formation of immune complexes may advance immunopathology. Enhancement of infection due to uptake of antibody-opsonized virus by Fc receptor-bearing cells or, as recently described, by triggering of the spike by a distinct type of antibodies to the N-terminal domain (NTD), may occur (69). We observed no negative impact of plasma treatment in our study. None of the measured antibody parameters was associated with slowing of viral clearance. The overall mortality rate in the 30 treated participants was low (3.3%) and well below the 9-13% average mortality among hospitalized individuals in Switzerland (38, 39). This is in stark contrast to the CONCOR-1 convalescent plasma trial that reported a disease-exacerbating effect that was linked to non-neutralizing antibodies directed against the spike protein (26). The nature of the disease worsening effect was not resolved in CONCOR-1, but notably, participants portrayed by a high rate of severe disease (intubation or death by day 30) both in the treatment (32.4%) and standard of care arm (28%), as opposed to only 6.6% in our trial in which critical ill and already intubated patients could not be included. The equally comparatively high rates of serious adverse events (33.4%) and transfusion-related events (5.7%) after 500ml plasma treatment in CONCOR-1 highlight the population- and procedure-related differences that generally affect outcome analyses of convalescent plasma studies. To improve tolerability, our study design foresaw three smaller sized plasma units (200 ml) to be given on three consecutive days to patients who had no signs of circulatory instability nor ARDS. With this selection of recipients, we observed a safety profile of CP that is comparable to the one of plasma administered for other indications. Another notable difference of our trial to CONCOR-1 and other convalescent plasma trials was that, according to Swiss regulations in transfusion medicine (70), exclusively male donors were allowed, in order to exclude TRALI reaction after transfusion of plasma obtained from female donors. While our study is comparatively small, the absence of transfusion-related adverse events, supports that low volume application together with restriction to male plasma donation should be generally considered to limit adverse events by convalescent plasma therapy.

We consider several factors to be crucial as to why our study showed an effect of convalescent plasma therapy that was not observed in other studies. First, the recipients were at an early stage of infection and received the plasma therapy before the peak of endogenous antibody activity, allowing the passively administered antibodies to exert their effect. Second, the convalescent plasma was not selected for antibody content, which provided the opportunity to perform an effective component analysis to decipher if and which antibody parameters in donor plasma have an impact. Third, multifactorial profiling of neutralizing and binding antibody responses proved critical. Fourth, our study highlights the importance of assessing the immune status of patients when analyzing SARS-CoV-2 therapies. As we show, systemic viremia is inversely related to the presence of endogenous neutralizing antibody activity and should be considered a relevant marker of immunity that is not yet mature, delayed or impaired. Our study shows a strong effect of endogenous neutralizing antibody on viral clearance, which is most pronounced in individuals who had not developed neutralizing activity at baseline. Taken together, these data argue that beyond antibody-based interventions, SARS-CoV-2 therapeutics in general must be evaluated in the context of the dynamic state of the endogenous response, as both evolving endogenous immunity and the therapeutic agent will have an impact on viral clearance.

## Methods

### Trial design and ethical approval

Convalescent plasma therapy is not approved for COVID-19 patients in Switzerland, the study was therefore classified by the responsible Swiss authorities for drugs and medical products, Swissmedic (www.swissmedic.ch) as first in human trial, under the category C (Swissmedic 2020TpP1004). Therefore, no placebo arm was allowed and tolerability was addressed as a primary objective. We accordingly designed this phase I clinical trial as an open-label, nonrandomized, single-center clinical trial (ClinicalTrials.gov Identifier: NCT04869072) to evaluate the safety (primary outcome) and potential efficacy (secondary outcome) of SARS-CoV-2 convalescent plasma in COVID-19 disease (Fig.1A, Table S1-S4). The study focused on individuals with advanced COVID-19 disease that required hospitalization but not intensive care. The study was initiated in April 2020 and completed in November 2020 and was approved by the local ethics committee of the Canton of Zurich, Switzerland (BASEC 2020-00787). Written informed consent was obtained from all study participants and convalescent plasma donors. In brief, included patients received three units of single-donor plasma donation (200 ml per unit) on three consecutive days, followed by an observation period of 70 days, during which clinical and laboratory parameters were monitored. Data collection and monitoring was coordinated by the Clinical Trials Center (CTC), located at the trial site (University Hospital Zurich) and included an internet-based secure data base secuTrial® for data and query management, monitoring, reporting and coding. An independent data monitoring committee (IDMC) received weekly reports to assess safety and decide on continuation or termination of the trial.

### Study participants: inclusion and exclusion criteria

Patients hospitalized with COVID-19 were recruited at a single trial site (University Hospital Zurich) prior to availability of vaccines and monoclonal antibody therapeutics in Switzerland. There was no time limit after SARS-CoV-2 diagnosis or onset of symptoms set for inclusion. Inclusion criteria were (1) signed informed consent; (2) COVID-19 diagnosis based on SARS-CoV-2 reverse transcription polymerase chain reaction (RT-PCR) testing; (3) aged at least 50 years and presence at least one of the following risk factors for a poor outcome: pre-existing cardiovascular disease, diabetic disease, chronic obstructive pulmonary disease (COPD), chronic liver disease, chronic renal failure, immunodeficiency, immunosuppression or neoplastic disease; or (4) aged at least 18 years and immunosuppressed or cancer (5); or aged at least 18 years and presence of at least one of the following signs of severe COVID19: SpO2 ≤ 94% on room air, O2-supplementation and typical changes on chest x-ray and/or lung-CT scan. COVID19-related exclusion criteria were the following, (1) life-threatening COVID-19 defined as hospitalization in the intensive care unit (ICU) or mechanical ventilation; (2) signs of acute respiratory distress syndrome (ARDS); (3) cytokine release syndrome. Patients with a known history of IgA deficiency were also excluded from the study.

### SARS-CoV-2 convalescent plasma

Fully recovered male individuals with previous PCR-confirmed SARS-CoV-2 infection, were recruited through the Zurich blood donation service as convalescent plasma donors. Plasma collection was performed after complete recovery and clearance of SARS-CoV-2, which was verified by RT-PCR testing in NPS before plasma collection. Donor screening and selection was based on the following criteria: male, aged above 18 years, fulfilling Swiss criteria for blood donation (70, 71), RT-PCR confirmed SARS-CoV-2 infection, two negative RT-PCR SARS-CoV-2 test results from nasopharyngeal swabs (at least 24 hours apart) before plasmapheresis. Convalescent plasma collection was performed based on routine plasma collection procedures via plasmapheresis technology, processed and pathogen inactivated using Intercept® technology according to standard operating procedures as approved by Swissmedic (72).Labelling of final product and release for transfusion were compliant to the requirements of the Federal Act on Medicinal Products and Medical Devices, Swissmedic and the prescription of Blutspende Schweiz. Each plasma donation was split into three aliquots containing 200 ml. The plasma products were immediately frozen (fresh-frozen plasma, FFP) and thawed upon use. In total, 75 donor plasma were collected throughout the trial period to ascertain the availability of ABO compatible plasma.

### Convalescent plasma transfusion

Three units of 200ml of convalescent plasma (FFP) of a single donor were transfused to one AB0 compatible recipient at three consecutive days (day 0, 1, 2; Fig. 1A). Convalescent plasma transfusion was administered at a rate of 100ml/hour. Paracetamol 1g and Clemastin 1mg were administered as premedication one hour before convalescent plasma transfusion. After the treatment period, plasma recipients were followed up for ten weeks, with safety visits performed two and seven days as well as three and ten weeks after the last plasma donation (day 2).

### Standard COVID-19 treatment

Patients received in addition to the convalescent plasma therapy the standard treatment for COVID-19 recommended at the time. Treatment consisted of symptomatic control and supportive care and was continuously adjusted to guidelines of the Swiss Society of Infectious diseases (73). Oxygen therapy was supplied as per requirement in all patients. The trial participants had access to all upcoming novel treatment options (e.g. remdesivir) during the trial. Depending on the availability of new therapeutics and treatment recommendations at the time of inclusion and the clinical indication, antiviral drugs (remdesivir) and steroids (dexamethasone) were provided in addition to individually required medications for COVID-19 unrelated causes including antibacterial therapy.

### Monitoring of transfusion related adverse events

Patients were carefully monitored by expert hematologists for occurrence of adverse events of plasma therapy including transfusion related acute lung injury (TRALI), anaphylactic reactions (AR), hemolytic reactions (HR) and transfusion associated circulatory overload (TACO).

### Clinical laboratory parameters

To assess safety and beneficial effects of convalescent plasma therapy as defined in the trial’s outcome measures (Table S4) an extended evaluation of clinical parameters was conducted based on routine diagnostic analyses from chemistry, hematology, immunology, microbiology and virology (Table S3). SARS-CoV-2 specific analyses are detailed below.

### Assessing overall clinical status

We included 7-category “pulmonary” ordinal outcome used in the TICO trial as a measure to rate overall clinical improvement as described (43). This pulmonary outcome allows a longitudinal assessment of disease stage and thus provides a more detailed information compared to time to hospital discharge, a commonly used easy accessible parameter of improvement. Importantly, during the initial phase of the pandemic, the tense situation in local nursing homes and recovery facilities often delayed the discharge of patients from the hospital to home care immediately after COVID-19 recovery in the study hospital. As thus length of hospitalization must be viewed as partially confounded, we did not include it as a parameter in sub-analyses.

### Quantitative SARS-CoV-2 RT-PCR

Nasopharyngeal swabs and plasma were analyzed for SARS-CoV-2 by RT PCR utilizing the Cobas SARS-CoV-2 IVD test (Roche) as described (74). Samples are recorded negative, when both E-gene and ORF-1 are not detected. Ct values are recorded for positive RT-PCR results.

### ABCORA 2 multiplex SARS-CoV-2 serology profiling

Seroreactivity to SARS-CoV-2 was profiled in EDTA plasma using the in-house developed multiplex bead assay ABCORA 2 (50). The assay measures IgG, IgA and IgM reactivity to four SARS-CoV-2 antigens RBD, S1, S2 and N (12 SARS-CoV-2 parameters) in addition to IgG, IgA and IgM reactivity to S1 of HCoV-HKU1. Briefly, pre-diluted plasma (1/100) was incubated with antigen loaded MagPlex beads (Luminex Corporation, Austin, TX) with unique fluorescent bead regions. Bound immunoglobulin was detected with phycoerythrin (PE)-labeled detector antibodies for IgG, IgA or IgM. SARS-CoV-2 positive plasma reactivity was defined using the ABCORA 2.3 computational approach achieving 99.07% specificity and 94.29% sensitivity (50). Antibody binding titers were defined by serial plasma dilution and recorded as 50% effective titer concentrations (EC50) using a four-parameter logistic curve (y=Bottom+(Top-Bottom)/(1+10^((logEC50-X)*HillSlope) as described.

### Elecsys® Anti-SARS-CoV-2 S assay

Donor plasma were evaluated with the Elecsys® Anti-SARS-CoV-2 S assay (Roche Diagnostics GmbH), a quantitative ECLIA immunoassays that detects antibodies to the SARS-CoV-2 S protein RBD, according to the manufacturers instruction. Results are recorded in U/mL. Values ≥0.80 U/mL are recorded as positive for anti-SARS-CoV-2 S antibodies.

### SARS-CoV-2 pseudo-neutralization assay

SARS-CoV-2 plasma neutralization activity was assessed using a HIV-based pseudovirus system as described (50, 75). Particles of the env-inactivated HIV-1 reporter construct pHIV-1NL4-3 ΔEnv-NanoLuc (pHIV-1Nanoluc; provided by P. Bieniasz, Rockefeller University, NY, USA) were pseudotyped with codon optimized, truncated SARS-CoV-2 spike (expression plasmid P_CoV2_Wuhan) by co-expression in HEK 293-T cells. HEK 293-T cells were obtained from the American Type Culture Collection. Infection of HT1080/ACE2cl.14 cells (provided by P. Bieniasz, Rockefeller University, NY, USA) with SARS-CoV-2 pseudoparticles was detected by measuring the NanoLuc luciferase reporter activity in cell lysates 48 h post infection using the Nano-Glo Luciferase Assay System (Promega, Fitchburg, WI) and readout on a Perkin Elmer EnVision reader. Neutralization tests of 1/100 diluted plasma were conducted in 384-well plates as described (50). Plasma neutralization titers causing 50% reduction in viral infectivity (NT50) compared to controls without plasma were calculated by fitting a sigmoid dose–response curve (variable slope) to the RLU data, using GraphPad Prism with constraints (bottom=0, top=100). If 50% inhibition was not achieved at the lowest plasma dilution of 1/100, a ’less than’ value was recorded. All measurements were conducted in duplicates.

### SARS-CoV-2 whole genome sequencing

SARS-CoV-2 whole-genome sequencing was performed according to the nCoV-2019 sequencing protocol v3 (LoCost) V.3 (76, 77). Briefly, total nucleic acids were extracted followed by reverse transcription with random hexamers using LunaScript RT SuperMix Kit (NEB). The generated cDNA was used as input for two pools of overlapping PCR reactions (ca. 400nt each) spanning the viral genome using Q5 Hot Start High-Fidelity 2X Master Mix (NEB). Amplicons were pooled per patient before NexteraXT library preparation and sequencing on an Illumina MiSeq for 1 × 151 cycles. To generate SARS-CoV-2 consensus sequences, reads were iteratively aligned using SmaltAlign (78).

### Outcome and statistical analysis

Statistical analyses were performed in R (Version 4.0.5). Figures were made using the ggplot2 package (79). All reported statistical analyses are of explorative, descriptive nature. We therefore opted not to adjust for multiple testing.

Completion of plasma therapy (day 2, after third dose) was defined as End of Treatment (EOT). To assess benefits of treatment, laboratory and respiratory outcomes were compared between baseline and follow-up visits using a paired t-test. Plasma recipients with missing values either at baseline or after therapy were not considered in this analysis.

Analyzing the difference of clinical, laboratory and immunologic parameters between different groups among recipients (e.g. between recipients receiving plasma with high or low neutralization titers) was done by unpaired t-test. Differences in times to viral clearance according to given subgroups were assessed either by a Kaplan-Meier analysis or by interval-censored survival parametric survival models. In the Kaplan-Meier analyses time to viral clearance was defined as the first negative PCR test (no Ct or Ct ≥ 45) that was not followed by any positive test.

Our parametric survival model assumed a Gamma-distributed time to clearance and used the last positive and first negative SARS-CoV-2 RT-PCR tests in NPS to define the interval in which the virus was cleared. The difference in time to viral clearance between the two groups was modelled assuming proportional hazard. We adjusted these analyses for two potential confounders: baseline viral load and the presence of any comorbidity. Finally, we determined the impact of the donor plasma on virus decay using censored regression.

## Data Availability

All data produced in the present study are available upon reasonable request to the authors

## Acknowledgments

General

## Author contributions

Each author’s contribution(s) to the paper should be listed [we encourage you to follow the CRediT model]. Each CRediT role should have its own line, and there should not be any punctuation in the initials.

Conceptualization and study design: HFG, MGM, AT, BF

Clinical investigation: MM, DLB, EW, RDK, HFG, ES, BF, MGM, EW

Laboratory investigation: IAA, SE, AA, MH, SS, MZ, MS, JW, PR, JB, VK

Methodology: AT, RDK, IAA

Statistics: AH, RDK, CP

Visualization: MS, IAA

Funding acquisition: HFG, MGM, AT, BF

Study monitoring: MM, EW, RK

Supervision: AT, IAA, RDK, MGM, BF

Writing – original draft: AT, IAA, MM

Writing – review & editing: MM, IAA, AH, RK, MS, DLB, SE, AA, JW, PR, ES, CP, EW, JB, VK, MH, MZ, SS, BF, RDK, HFG, MGM, AA

## Data and materials availability

All available trial related data are available in the main text or the supplementary materials.

## Supplementary Materials for

**Table S1:** Study schedule

**Table S2**: Inclusion/Exclusion criteria of trial participants (plasma recipients)

**Table S3:** Monitored parameters

**Table S4**. Primary and secondary outcomes

**Table S5**. Overview comorbidities

**Table S6**: Safety outcomes

**Table S7**: Severe adverse events (SAE)

**Table S8**: Pulmonary ordinal outcome

**Table S9**: Severity outcome of trial participants assessed by pulmonary ordinal outcome

**Table S10**: Laboratory measures and clinical parameters

**Table S11**: Summary convalescent donor demographics

**Table S12:** Hazard ratios of time to viral clearance for binding and neutralizing parameters, presence of comorbidity and baseline viral load in nasopharyngeal swab.

**Fig. S1:** Longitudinal assessment of primary and secondary outcomes

**Fig. S2:** Phylogenetic analysis of the SARS-CoV-2 isolates from trial participants

**Fig. S3:** No impact of neutralization activity on hospital release

**Fig. S4:** Sensitivity analysis for impact of plasma on viral clearance

**Fig. S5:** Spike specific binding and neutralizing antibodies are linked with rapid virus clearance

**Fig. S6:** No impact on viral clearance when stratifying based on the Elecsys assay

**Fig. S7:** Sensitivity of time to viral clearance when removing patients on remdesivir

**Fig. S8:** High neutralizing plasma leads to faster virus decay in nasopharyngeal swabs

**Fig. S9:** SARS-CoV-2 antibody reactivities are correlated within antibody classes

**Fig. S10:** Evolution of SARS-CoV-2 antibodies in trial participants

**Fig. S11:** Antibody response over time in the three trial participants incapable of mounting antibody response

**Fig. S12:** Sensitivity survival analysis adjusting for the impact of endogenous neutralization on virus clearance

**Fig. S13:** Sensitivity survival analysis adjusting for the impact of endogenous binding antibodies on virus clearance.

