## Supplementary Material for "Contribution of endogenous and exogenous antibodies to clearance of SARS-CoV-2 during convalescent plasma therapy"

*Marconato et al*.

**Summary of supplementary tables and figures**

**Table S1:** Study schedule

**Table S2**: Inclusion/Exclusion criteria of trial participants (plasma recipients)

**Table S3:** Monitored parameters

**Table S4.** Primary and secondary outcomes

**Table S5.** Overview comorbidities

**Table S6**: Safety outcomes

**Table S7**: Severe adverse events (SAE)

**Table S8**: Pulmonary ordinal outcome

**Table S9**: Severity outcome of trial participants assessed by pulmonary ordinal outcome

**Table S10**: Laboratory measures and clinical parameters

**Table S11**: Summary convalescent donor demographics

**Table S12:** Hazard ratios of time to viral clearance for binding and neutralizing parameters, presence of comorbidity and baseline viral load in nasopharyngeal swab.

**Fig. S1:** Longitudinal assessment of primary and secondary outcomes

**Fig. S2:** Phylogenetic analysis of the SARS-CoV-2 isolates from trial participants

**Fig. S3:** No impact of neutralization activity on hospital release

**Fig. S4:** Sensitivity analysis for impact of plasma on viral clearance

**Fig. S5:** Spike specific binding and neutralizing antibodies are linked with rapid virus clearance

**Fig. S6:** No impact on viral clearance when stratifying based on the Elecsys assay

**Fig. S7:** Sensitivity of time to viral clearance when removing patients on remdesivir

**Fig. S8:** High neutralizing plasma leads to faster virus decay in nasopharyngeal swabs

**Fig. S9:** SARS-CoV-2 antibody reactivities are correlated within antibody classes

**Fig. S10:** Evolution of SARS-CoV-2 antibodies in trial participants

**Fig. S11:** Antibody response over time in the three trial participants incapable of mounting antibody response

**Fig. S12:** Sensitivity survival analysis adjusting for the impact of endogenous neutralization on virus clearance

**Fig. S13:** Sensitivity survival analysis adjusting for the impact of endogenous binding antibodies on virus clearance.

**Table S1:** Study schedule

| **Study period** | **Screening** | **Treatment period** | | | | | **Safety follow-up** | |
|---|---|---|---|---|---|---|---|---|
| **Visit** | **1** | **2** | **3** | **4** | **5** | **6** | **7** | **8** |
| **Time** (days) | **0** | **0** | **1** | **2** | **4** | **9** | **23+/-1** | **72+/-1** |
| Patient information and informed consent | **x** | | | | | | | |
| In-/exclusion criteria | **x** | **x** | | | | | | |
| Medical history and demographic data | **x** | | | | | | | |
| Documentation of concomitant drugs | **x** | **x** | **x** | **x** | **x** | **x** | **x** | **x** |
| Body weight/height | **x** | **x** | | | | | **x** | **x** |
| Physical examination | **x** | | | | | | **x** | **x** |
| Vital signs | **x** | **x** | **x** | **x** | **x** | **x** | **x** | **x** |
| **Screening laboratory measurements** | **x** | | | | | | | |
| **Blood typing** | **x** | | | | | | | |
| **Safety laboratory tests** | **x** | **x** | **x** | **x** | **x** | **x** | **x** | **x** |
| **Laboratory measures** | **x** | **x** | **x** | **x** | **x** | **x** | **x** | **x** |
| Urine pregnancy test | **x** | | | | | | | **x** |
| Samples for biobank (optional) | **x** | | | | **x** | **x** | **x** | **x** |
| Administer study medication | | **x** | **x** | **x** | | | | |
| Severe adverse/adverse events | **x** | **x** | **x** | **x** | **x** | **x** | **x** | **x** |

**Table S2**: **Inclusion/exclusion criteria of trial participants**

| **Inclusion** criteria: | **Exclusion** criteria: |
|---|---|
| A) Proven SARS-CoV-2 by PCR and hospitalization for COVID-19 in combination with either (1) or (2):<br>(1) Age ≥ 50<br><br>AND (at least one):<br>● Pre-existing cardiovascular disease<br>● Diabetic disease<br>● Immunodeficiency/immunosuppression<br>● Neoplastic disease<br>● COPD or chronic liver disease or chronic renal failure<br><br>(2) Age ≥ 18<br><br>AND (at least one):<br>● SpO2 ≤ 94% on room air or requiring supplemental oxygen at screening<br>● Typical changes on chest x-ray and/or lung-CT scan<br>● Immunosuppression or neoplastic disease<br><br>B) Written informed consent as documented by signature (Appendix Informed Consent Form) of the patient or, in case of inability, of the next relative/care-taking person. In the latter case, an independent doctor will also be involved and her/his signature will be required in order to enrol the patient. | 1. Contraindications to the class of drugs under study, e.g. known hypersensitivity or allergy to class of drugs or the investigational product (FFP)<br>2. Known IgA deficiency<br>3. Cytokine Release Syndrome grade ≥ 3<br>4. Acute respiratory distress syndrome (ARDS)<br>5. Patients already hospitalized in intensive care unit and/or already receiving mechanical ventilation<br>6. Known or suspected non-compliance, drug or alcohol abuse<br>7. Previous enrolment into the current study<br>8. Enrolment of the investigator, his/her family members, employees and other dependent persons<br>9. Women who are pregnant or breast feeding<br>10. Intention to become pregnant during the course of the study<br>11. Lack of safe contraception, defined as: Female participants of childbearing potential, not using and not willing to continue using a medically reliable method of contraception for the entire study duration, such as oral, injectable, or implantable contraceptives, or intrauterine contraceptive devices, or who are not using any other method considered sufficiently reliable by the investigator in individual cases. Please note that female participants who are surgically sterilised/hysterectomised or post-menopausal for longer than 2 years are not considered as being of child bearing potential |

**Table S3: Monitored parameters**

| Monitored clinical parameter and laboratory measures | Baseline | Treatment period | Follow-up period |
|---|---|---|---|
| | Day 0 | Day 0-2, 9 | 23 +/-1, 72+/-1 |
| **Clinical parameters:** | | | |
| Blood pressure (mmHg), Heart rate (bpm*)<br>Respiratory rate (bpm**)<br>Body temperature (°C, ear)<br>Oxygen saturation (SpO2 (%)) | X | X | X |
| **Laboratory measures:** | | | |
| Blood count (WBC subsets), INR<br>CRP (mg/L), Ferritin (ug/L), LDH (unit/L)<br>D-dimer (mg/L), Fibrinogen (g/L), IL-6<br>Na, K, Ca, , Serum Creatinine (mg/dL)<br>Liver enzymes: AST (SGOT) (U/L), ALT (SGPT) (U/L), γGT, Troponin I, Myoglobin<br>Rapid urine test | X | X | X |
| 25-OH Vitamin, Albumin<br>Urin sediment<br>IgG, IgG subclasses, IgA, IgM<br>PT, PTT | X | | |
| SARS-CoV-2 antibody binding response (IgM, IgG, IgA response against S1, S2, RBD, N) | X | | X |
| SARS-CoV-2 neutralizing antibody binding titers (NT50) | X | | |
| PCR for SARS-CoV-2 on nasal swab, plasma, BAL | X | X | X |

*beat per minute, **breaths per minute,

**Table S4. Primary and secondary outcomes**

| |
|---|
| **Primary outcomes:** |
| **(A) Safety measures** |
| **1. Assessment for**<br>• Transfusion related acute lung injury (TRALI)<br>• Anaphylactic reactions<br>• Hemolytic reactions<br>**2. Clinical monitoring and measurement of laboratory parameters**<br>• During treatment episode: Day 0,1,2<br>• Follow up : Day 4, Day 9, Day 23 +/- 1 and Day 72 +/- 1 |
| **(B) Efficacy measures** |
| **1. Improvement of respiration:**<br>• Respiratory rate (breaths per minute, bpm), Oxygen saturation (SpO2 (%))<br>**2. Improvement of laboratory measures:**<br>• CRP (mg/L),<br>• Ferritin (ug/L)<br>• lL-6 (mg/L)<br>• D-dimer (mg/L)<br>• Fibrinogen (g/L)<br>• LDH (unit/L)<br>**3. Reduction of hospitalization duration**<br>**4. Prevention of ICU-admission or shortening of ICU-hospitalization** |
| **Secondary outcomes:** |
| **1. Characterize humoral response (binding and neutralization) against SARS-CoV-2**<br>**2. Longitudinally assess viral shedding in nasopharyngeal swabs and blood samples** |

#### Table S5. Overview comorbidities

| ID | Gender | Comorbidity | Comorbidity type | Immune suppression | Impaired antibody response | Endogenous SARS-CoV-2 neutralization at baseline | Log10 viral load NPS at baseline | Viral suppression blood at baseline | Remdesivir treatment | Steroid treatment | Convalescent plasma (NT50 low/high) | Clinical outcome at study completion (day 72) |
|---|---|---|---|---|---|---|---|---|---|---|---|---|
| 1 | Female | Yes | Solid organ transplant (kidney) | yes | No | Yes | 4.7 | Yes | No | No | high | 1 |
| 2 | Male | Yes | Cardiovascular disease | No | Yes | No | 3.93 | Yes | No | No | high | 2 |
| 3 | Female | Yes | Cardiovascular disease, Diabetic disease | No | No | Yes | 4.48 | Yes | No | No | high | 1 |
| 4 | Male | Yes | Cardiovascular disease, Neoplastic disease (Diffuse large-cell B cell NHL) | Yes | Yes | No | 5.01 | No | No | No | high | 1 |
| 5 | Male | Yes | Cardiovascular disease | No | No | Yes | 4.34 | Yes | No | No | low | 1 |
| 6 | Female | No | | No | No | Yes | 3.54 | Yes | No | No | low | 1 |
| 7 | Male | Yes | Cardiovascular disease | No | No | Yes | 4.65 | Yes | No | No | low | 1 |
| 8 | Male | No | | No | No | Yes | 4.62 | Yes | No | No | high | 1 |
| 9 | Female | No | | No | No | Yes | 5.32 | No | No | No | high | 1 |
| 10 | Female | Yes | Cardiovascular disease, Diabetic disease | no | No | No | 2.81 | No | No | Yes | high | 1 |
| 11 | Male | Yes | Cardiovascular disease, Neoplastic disease (plasma cell leukaemia) | Yes | No | Yes | 4.43 | Yes | Yes | No | high | 1 |
| 12 | Male | Yes | Cardiovascular disease, Diabetic disease, COPD | No | No | No | 5.86 | No | No | No | low | 1 |
| 13 | Male | Yes | Cardiovascular disease | No | No | Yes | 4.79 | Yes | No | No | low | 1 |
| 14 | Male | No | | No | No | Yes | 3.77 | No | No | No | high | 1 |
| 15 | Female | Yes | Cardiovascular disease, Neoplastic disease (CLL) | Yes | Yes | No | 9.85 | No | Yes | No | high | 7 |
| 16 | Male | Yes | Cardiovascular disease, Diabetic disease | No | No | Yes | 3.91 | No | No | No | low | 1 |
| 17 | Female | Yes | Cardiovascular disease, Diabetic disease | No | No | Yes | 3.79 | Yes | No | No | low | 1 |
| 18 | Female | No | | No | No | No | 5.8 | No | Yes | No | low | 2 |
| 19 | Male | Yes | Cardiovascular disease, COPD | No | No | No | 6.42 | No | No | No | low | 1 |
| 20 | Male | No | | No | No | No | 3.85 | No | No | No | low | 1 |
| 21 | Female | Yes | Cardiovascular disease, Diabetic disease | No | No | Na | 4.39 | Yes | No | No | high | 1 |
| 22 | Male | Yes | Cardiovascular disease, Chronic renal failure | No | No | Yes | 5.15 | Yes | No | No | high | 1 |
| 23 | Male | Yes | Cardiovascular disease, Diabetic disease | No | No | No | 3.15 | No | No | No | high | 1 |
| 24 | Male | Yes | Cardiovascular disease | No | No | Yes | 6.97 | No | No | Yes | high | 2 |
| 25 | Male | Yes | Cardiovascular disease | No | No | Yes | 4.4 | No | No | No | high | 1 |
| 26 | Male | Yes | Cardiovascular disease | No | No | No | 5.56 | No | No | Yes | low | 2 |
| 27 | Male | No | | No | No | Yes | 3.57 | Yes | No | No | low | 1 |
| 28 | Female | Yes | Cardiovascular disease, Diabetic disease | No | No | Yes | 5.19 | No | No | Yes | high | 1 |
| 29 | Male | No | | No | No | No | 4.35 | No | Yes | Yes | low | 1 |
| 30 | Male | Yes | Solid organ transplant (kidney) | Yes | No | No | 4.56 | Yes | No | No | high | 1 |

**Table S6**: **Safety outcomes**

| | Total |
|---|---|
| **Complications of CPT therapy (safety), No (%)** | |
| Transfusion Related Acute Lung Injury (TRALI) | 0 (0%) |
| Anaphylactic Reactions (AR) | 0 (0%) |
| Hemolytic Reactions (HR) | 0 (0%) |
| Transfusion associated circulatory overload (TACO) | 0 (0%) |
| **Clinical and laboratory outcomes (efficacy), No (%)** | |
| COVID-19 related death during the study | 0 (0%) |
| Hospitalization in ICU | 2/30 (6.67%) |
| Mechanical ventilation | 2/30 (6.67%) |

**Table S7**: **Severe adverse events (SAE)**

| ID | Gender | Description of event | Description of intervention | Outcome | Comments, if relevant |
|---|---|---|---|---|---|
| 3 | Female | NSTEMI | No specific treatment needed | improved | Possible correlation with COVID-19 |
| 10 | Female | Hospital acquired pneumonia (HAP) | antibiotics | resolved | COVID-19 resolved, HAP |
| 15 | Female | Hospital acquired pneumonia (HAP) | antibiotics | death | COVID-19 resolved, HAP |
| 17 | Female | Pulmonary Embolism | Anticoagulation with heparin and liquemin | resolved | Complication of COVID-19 |
| 18 | Female | Respiratory insufficiency due to COVID-19 | Patient transferred to the ICU and mechanical ventilation needed | resolved | - |
| 20 | Male | Arthritis (right lateral malleolus) | Local corticosteroids | resolved | - |
| 26 | Male | Respiratory insufficiency due to COVID-19 | Patient transferred to the ICU and mechanical ventilation needed | resolved | |

**Table S8**: **Pulmonary ordinal outcome**

| |
|---|
| 1= Can independently undertake usual activities with minimal or no symptoms |
| 2= Symptomatic and currently unable to independently undertake usual activities but no need of supplemental oxygen (or not above premorbid requirements) |
| 3= Supplemental oxygen (<4 liters/min, or <4 liters/min above premorbid requirements) |
| 4= Supplemental oxygen (≥4 liters/min, or ≥4 liters/min above premorbid requirements, but not high-flow oxygen) |
| 5= Non-invasive ventilation or high-flow oxygen |
| 6=Invasive ventilation, extracorporeal membrane oxygenation (ECMO), mechanical circulatory support, or new receipt of renal replacement therapy |
| 7=Death |

**Table S9**: **Severity outcome of study participants assessed by pulmonary ordinal outcome**

| | Before/during therapy | | | Follow-up visits | | Safety visits | |
|---|---|---|---|---|---|---|---|
| | Day 0 | Day 1 | Day 2 | Day 4 | Day 9 | Day 23 +/- 1 | Day 72 +/- 1 |
| **Ordinal scale** | | | | | | | |
| 1 | 0 (0%) | 0 (0%) | 0 (0%) | 1 (3%) | 12 (40%) | 21 (70%) | 25 (83%) |
| 2 | 14 (47%) | 16 (53%) | 14 (47%) | 16 (53%) | 12 (40%) | 6 (20%) | 4 (13%) |
| 3 | 10 (33%) | 9 (30%) | 10 (33%) | 9 (30%) | 3 (10%) | 1 (3%) | 0 (0%) |
| 4 | 5 (17%) | 4 (13%) | 4 (13%) | 2 (7%) | 1 (3%) | 0 (0%) | 0 (0%) |
| 6 | 1 (3%) | 1 (3%) | 2 (7%) | 2 (7%) | 2 (7%) | 1 (3%) | 0 (0%) |
| 7 | 0 (0%) | 0 (0%) | 0 (0%) | 0 (0%) | 0 (0%) | 1 (3%) | 1 (3%) |

**Table S10**: **Longitudinal laboratory measures and clinical parameters of study participants**

| | **Day 0** | **Day 1** | | **Day2** | | **Day 4** | | **Day 9** | | **Day 23 +/- 1** | | **Day 72 +/- 1** | |
|---|---|---|---|---|---|---|---|---|---|---|---|---|---|
| **Clinical parameters** | median (IQR) | median (IQR) | p-value | median (IQR) | p-value | median (IQR) | p-value | median (IQR) | p-value | median (IQR) | p-value | median (IQR) | p-value |
| Respiratory rate | 18 (16-22) | 18 (16.2-24) | 0.56 | 18 (16.2-23.8) | 0.4 | 18 (14-22) | 0.12 | 14 (14-17.5) | 0.0015 | 14 (14-14) | <0.001 | 14 (12-14) | <0.001 |
| Oxygen saturation level | 94 (93-96) | 94 (93-95) | 0.56 | 94 (92.2-96) | 0.45 | 95 (94-96) | 0.36 | 97 (96-98) | <0.001 | 97 (96-98) | <0.001 | 97 (97-98) | <0.001 |
| Temperature (in °C) | 37.1 (36.8-37.5) | 37.1 (36.8-37.6) | 0.85 | 37.1 (36.8-37.4) | 0.85 | 36.9 (36.6-37.2) | 0.043 | 36.8 (36.4-37) | 0.028 | 36.8 (36.5-36.9) | <0.001 | 36.6 (36.5-36.8) | <0.001 |
| **Laboratory parameters** | median (IQR) | median (IQR) | p-value | median (IQR) | p-value | median (IQR) | p-value | median (IQR) | p-value | median (IQR) | p-value | median (IQR) | p-value |
| CRP | 98.5 (66.2-143.8) | 92.5 (53.8-117.2) | 0.11 | 76.5 (42.2-99.2) | 0.0038 | 40 (19.2-58) | <0.001 | 6.4 (3.5-18.8) | <0.001 | 1.4 (0.8-4.9) | <0.001 | 1.5 (0.6-2.6) | <0.001 |
| D-dimer | 0.705 (0.5-1.2) | 0.64 (0.4-1) | 0.47 | 0.755 (0.5-1.1) | 0.72 | 0.75 (0.5-1.1) | 0.73 | 0.61 (0.4-1.4) | 0.46 | 0.53 (0.4-0.8) | 0.3 | 0.365 (0.3-0.5) | 0.012 |
| Ferritin | 691 (456.5-1219.8) | 716 (425.8-1152) | 0.86 | 768 (486-1389) | 0.92 | 734.5 (327.2-1278.2) | 0.75 | 454.5 (249.8-592) | 0.04 | 234 (122-311.5) | <0.001 | 74 (43-170.2) | <0.001 |
| Fibrinogen | 5.6 (5-6.1) | 5.1 (4.7-5.5) | 0.22 | 5.2 (4.7-6) | 0.31 | 4.9 (4.3-5.8) | 0.0062 | 4.1 (3.3-4.6) | <0.001 | 3.3 (2.9-3.8) | <0.001 | 3 (2.7-3.5) | <0.001 |
| IL-6 | 33.05 (18.6-53) | 35.7 (21.2-67.3) | 0.7 | 31 (14-49.7) | 0.59 | 10.6 (4.8-26.6) | 0.22 | 4.7 (3.3-10.3) | 0.033 | 2.7 (1.9-4.1) | 0.027 | 2.4 (1.9-4) | 0.036 |
| LDH | 547.5 (489.5-687.5) | 592 (522.8-713.8) | 0.64 | 587 (504-703) | 0.83 | 550 (459-696) | 0.74 | 477 (410.8-545.8) | 0.01 | 368 (326-458) | <0.001 | 401.5 (358.8-439.8) | <0.001 |

**Table S11**: **Summary convalescent donor demographics**

| | **Convalescent plasma (banked)** (n=75) | **Transfused** (n=30) | | | |
|---|---|---|---|---|---|
| | | Transfused (total) | NT50<250 | NT50>250 | p-value |
| **Age**, median (IQR) | 33 (21-45.5) | 26.5 (20-38.5) | 26 (19-36) | 27 (20-39) | 0.73 |
| **Sex**, No. (%) | | | | | |
| Male | 75 (100%) | 30 (100%) | 13 (100%) | 17 (100%) | 1.00 |
| **Blood type**, No (%) | | | | | |
| A | 34 (45%) | 13 (43%) | 6 (46%) | 7 (41%) | 1 |
| B | 9 (12%) | 5 (17%) | 1 (8%) | 4 (24%) | 0.35 |
| AB | 1 (1%) | 1 (3%) | 1 (8%) | 0 (0%) | 0.43 |
| O | 28 (37%) | 11 (37%) | 5 (38%) | 6 (35%) | 1 |
| **Symptoms duration**, median (IQR) | 12 (8-16.5) | 10.5 (7-15.5) | 10 (7-14) | 11 (7-16) | 0.74 |

**Table S12:** Hazard ratios of time to viral clearance for binding and neutralizing parameters, presence of comorbidity and baseline viral load in nasopharyngeal swab. Estimates for both univariable and multivariable models are given.

| Antibody | Antigen | Binding/neutralizing antibody | | | | Baseline viral load | | | | Presence of comorbidity | | | |
|---|---|---|---|---|---|---|---|---|---|---|---|---|---|
| | | Univariate model | | Multivariate model | | Univariate model | | Multivariate model | | Univariate model | | Multivariate model | |
| | | HR (95% CI) | p-value | HR (95% CI) | p-value | HR (95% CI) | p-value | HR (95% CI) | p-value | HR (95% CI) | p-value | HR (95% CI) | p-value |
| Neutralization | | 2.4 [1,5.9] | 0.05 | 3 [1.1,8.1] | 0.026 | 0.53 [0.34,0.84] | 0.0064 | 0.52 [0.32,0.84] | 0.008 | 0.56 [0.22,1.4] | 0.23 | 0.62 [0.22,1.8] | 0.36 |
| Roche | | 2.5 [1,6] | 0.048 | 2.1 [0.75,5.6] | 0.16 | 0.53 [0.34,0.84] | 0.0064 | 0.46 [0.27,0.78] | 0.0036 | 0.56 [0.22,1.4] | 0.23 | 0.91 [0.33,2.5] | 0.85 |
| IgG | RBD | 2.7 [1.1,6.5] | 0.032 | 2.4 [0.93,6.2] | 0.071 | 0.53 [0.34,0.84] | 0.0064 | 0.5 [0.31,0.82] | 0.0055 | 0.56 [0.22,1.4] | 0.23 | 0.79 [0.28,2.2] | 0.66 |
| IgG | S1 | 1.5 [0.62,3.5] | 0.38 | 3.5 [1.3,9.3] | 0.013 | 0.53 [0.34,0.84] | 0.0064 | 0.5 [0.3,0.82] | 0.0061 | 0.56 [0.22,1.4] | 0.23 | 0.8 [0.28,2.3] | 0.68 |
| IgG | S2 | 1.5 [0.63,3.6] | 0.35 | 2.9 [1,7.9] | 0.042 | 0.53 [0.34,0.84] | 0.0064 | 0.46 [0.28,0.76] | 0.0024 | 0.56 [0.22,1.4] | 0.23 | 0.88 [0.32,2.5] | 0.81 |
| IgG | N | 1.6 [0.66,3.8] | 0.3 | 1.1 [0.44,2.6] | 0.89 | 0.53 [0.34,0.84] | 0.0064 | 0.54 [0.34,0.87] | 0.012 | 0.56 [0.22,1.4] | 0.23 | 0.92 [0.33,2.6] | 0.88 |
| IgG | RBD+S1+S2+N | 1.7 [0.72,4.1] | 0.22 | 2.5 [0.96,6.4] | 0.062 | 0.53 [0.34,0.84] | 0.0064 | 0.49 [0.3,0.8] | 0.0039 | 0.56 [0.22,1.4] | 0.23 | 0.85 [0.31,2.3] | 0.76 |
| IgA | RBD | 2.5 [1,6] | 0.048 | 6.1 [1.9,20] | 0.0027 | 0.53 [0.34,0.84] | 0.0064 | 0.4 [0.23,0.7] | 0.0011 | 0.56 [0.22,1.4] | 0.23 | 0.84 [0.3,2.4] | 0.74 |
| IgA | S1 | 1.7 [0.71,4.1] | 0.24 | 6.1 [1.9,20] | 0.0027 | 0.53 [0.34,0.84] | 0.0064 | 0.4 [0.23,0.7] | 0.0011 | 0.56 [0.22,1.4] | 0.23 | 0.84 [0.3,2.4] | 0.74 |
| IgA | S2 | 0.48 [0.19,1.2] | 0.1 | 4 [1.4,12] | 0.011 | 0.53 [0.34,0.84] | 0.0064 | 0.41 [0.24,0.69] | 0.00096 | 0.56 [0.22,1.4] | 0.23 | 0.73 [0.26,2.1] | 0.55 |
| IgA | N | 2.2 [0.88,5.2] | 0.092 | 5 [1.6,15] | 0.0049 | 0.53 [0.34,0.84] | 0.0064 | 0.4 [0.23,0.7] | 0.0015 | 0.56 [0.22,1.4] | 0.23 | 0.83 [0.3,2.3] | 0.72 |
| IgA | RBD+S1+S2+N | 0.95 [0.4,2.3] | 0.92 | 6.1 [1.9,20] | 0.0027 | 0.53 [0.34,0.84] | 0.0064 | 0.4 [0.23,0.7] | 0.0011 | 0.56 [0.22,1.4] | 0.23 | 0.84 [0.3,2.4] | 0.74 |
| IgM | RBD | 1.7 [0.71,4.1] | 0.23 | 2.5 [0.96,6.3] | 0.061 | 0.53 [0.34,0.84] | 0.0064 | 0.5 [0.31,0.83] | 0.0064 | 0.56 [0.22,1.4] | 0.23 | 0.73 [0.26,2] | 0.55 |
| IgM | S1 | 2.5 [1,6] | 0.048 | 4.1 [1.5,11] | 0.0063 | 0.53 [0.34,0.84] | 0.0064 | 0.45 [0.27,0.77] | 0.0031 | 0.56 [0.22,1.4] | 0.23 | 0.75 [0.27,2.1] | 0.58 |
| IgM | S2 | 1.7 [0.72,4.1] | 0.22 | 2.7 [1,7.2] | 0.049 | 0.53 [0.34,0.84] | 0.0064 | 0.46 [0.27,0.76] | 0.0026 | 0.56 [0.22,1.4] | 0.23 | 0.88 [0.32,2.5] | 0.81 |
| IgM | N | 2.8 [1.1,6.7] | 0.027 | 0.68 [0.26,1.8] | 0.45 | 0.53 [0.34,0.84] | 0.0064 | 0.57 [0.35,0.94] | 0.028 | 0.56 [0.22,1.4] | 0.23 | 0.98 [0.35,2.8] | 0.98 |
| IgM | RBD+S1+S2+N | 1.6 [0.69,4] | 0.26 | 3.4 [1.3,9.5] | 0.017 | 0.53 [0.34,0.84] | 0.0064 | 0.43 [0.25,0.72] | 0.0016 | 0.56 [0.22,1.4] | 0.23 | 0.97 [0.35,2.7] | 0.96 |

**Supplementary Figure legends**

**Fig. S1: Longitudinal assessment of primary and secondary outcomes**. (A) Longitudinal clinical assessment of efficacy measures of all trial participants (n=30). Respiratory rate (breaths per minute (bpm)), level of oxygen saturation (SpO2%) and body temperature (°C). (B) Longitudinal laboratory measurements as defined in primary outcomes of all trial participants (n=30): C-reactive protein (CRP), D-dimer, Ferritin, Fibrinogen, IL-6 and LDH. Levels of significance are assessed by paired t-test comparing baseline to subsequent measures. Only significant p-values were shown. Numbers of trial participants with available measurements are indicated.

**Fig. S2: Phylogenetic analysis of the SARS-CoV-2 isolates from 26 trial participants.** Phylogenetic analysis of sequences derived from the 28 SARS-CoV-2 sequences, together with the background SARS-CoV-2 sequences extracted from gisaid that represent viral diversity across the WHO Switzerland region Zurich during the same time frame. Wuhan/WH04/2020 (EPI_ISL_406801), belonging to clade 19B, was chosen as the outgroup. The SARS-CoV-2 lineages are shown in the figure with the patient's sequences labeled with numbers. The scale bar indicates the number of nucleotide substitutions per site.

**Fig. S3: No impact of neutralization activity on hospital release.** Kaplan-Meier analysis assessing the time to hospital release according to the neutralization level of the plasma received. High neutralization activity NT50>250 (purple), low neutralization activity NT50< 250 (light green). The log-rank test is used to calculate the p-value.

**Fig. S4**: **Sensitivity analysis for impact of plasma on viral clearance**. (A) Kaplan Meier analysis and (B) survival function estimated with parametric survival model for interval-censored data assessing the time to viral clearance according to neutralization level excluding three trial participants incapable of mounting antibody response (n=27). (C) and (D) corresponding analyses to (A) and (B) but restricting the population to trial participants who did not receive remdesivir (n=26). (E) and (F) corresponding analyses to (A) and (B) but restricting the population to trial participants who did not receive remdesivir and were capable of mounting antibody response (n=24).

**Fig. S5: Spike specific binding and neutralizing antibodies are linked with rapid virus clearance.** Kaplan-Meier analyses of the effect of the different antibody reactivities stratified by response levels for all combinations of antibody subtypes (IgG, IgA, IgA, IgM) and SARS-CoV-2 antigens (RBD, S1, S2 and NP) as well as the indicated sum of reactivities on the time to viral clearance. The effects of neutralization and binding activity as assessed with Roche S on the time to viral clearance are shown in the last row.

**Fig. S6: No impact on viral clearance when stratifying based on the Elecsys assay**. Forest plot depicting hazard ratios of univariable (grey) and multivariable (black) models of time to viral clearance including Elecsys Anti-SARS-CoV-2 (S) assay (U/ml). Multivariable analyses are corrected for baseline viral load and the presence of comorbidity. Low and high binding activity for each binding antibody parameter is stratified by the median binding reactivity.

**Fig. S7: Sensitivity of time to viral clearance when removing patients on remdesivir**. (A) Impact of antibody parameters on the time to viral clearance in multivariable parametric survival models when restricting to trial participants that did not receive remdesivir (n=26). Antibody reactivity of different antibody classes with SARS-CoV-2 antigens and neutralizing antibodies were included. Hazard ratios for individual antibody reactivities adjusted for the presence of comorbidity and the baseline viral load are shown. Significant results are marked in red. Low and high binding activity for each binding antibody parameter is stratified by the respective median binding reactivity. Low and high neutralization activity is stratified by a 50% neutralization titer of 250.

**Fig. S8: High neutralizing plasma leads to faster virus decay in nasopharyngeal swabs**. (A and B) Censored regression model estimating decay rate of viral load (log10 viral load) of nasopharyngeal swabs from time to treatment initiation when restricting to trial participants that did not receive remdesivir (n=26), according to (A) the level of neutralization (low neutralization NT50<250, light green; high neutralization, NT50>250, purple) or (B) level of binding defined by the ABCORA test sum of S1 SOC values. Low and high sum of S1 binding is stratified by the median binding reactivity. Significance was assessed using a two-sided t-test.

**Fig. S9**: **SARS-CoV-2 antibody reactivities are correlated within antibody classes.** Spearman correlation matrix assessing correlation between binding and neutralizing characteristics of the banked plasma samples. Levels of significance are assessed by asymptotic t approximation of Spearman's rank correlation. Color shading denotes correlation coefficient. Stars depict *p< 0.05, ** p< 0.01, *** p< 0.001.

**Fig. S10**: **Evolution of SARS-CoV-2 antibodies in trial participants**. Longitudinal binding antibody activity of trial participants (n=29) at baseline (day 0, purple), day 9 (green) and day 72 (yellow) assessed with the multiplex SARS-CoV-2 ABCORA test. Depicted are signal over cutoff (SOC) values of IgG, IgA, IgM against RBD and S1, S2 and N. Box plots depict the interquartile ranges with vertical lines representing the minimum and maximum values.

**Fig. S11**: **Antibody response over time in the three trial participants incapable of mounting antibody response**. Longitudinal binding antibody activity for the three trial participants assessed with the multiplex SARS-CoV-2 ABCORA test. Depicted are signal over cutoff (SOC) values of IgG, IgA, IgM against RBD, S1, S2 and N.

**Fig. S12**: **Sensitivity survival analysis adjusting for the impact of endogenous neutralization on virus clearance.** (A) Forest plot showing the hazard ratios from univariable (grey) and multivariable (black) models of time to viral clearance for baseline viral load, the presence of comorbidity and both the exogenous and endogenous neutralization, when restricting the population to trial participants that did not receive remdesivir (n=26) (B) and restricting to trial participants that were capable of mounting antibody response (n=27).

**Fig. S13: Sensitivity survival analysis adjusting for the impact of endogenous binding antibodies on virus clearance.** (A) Forest plot showing the hazard ratios from univariable (grey) and multivariable (black) models of time to viral clearance for baseline viral load, the presence of comorbidity and both the exogenous and endogenous S1 response (sum of IgG, IgA and IgM responses against S1), when restricting the population to trial participants that did not receive remdesivir (n=26) or (B) restricting to patients that were capable of mounting antibody response (n=26).

Figure S1

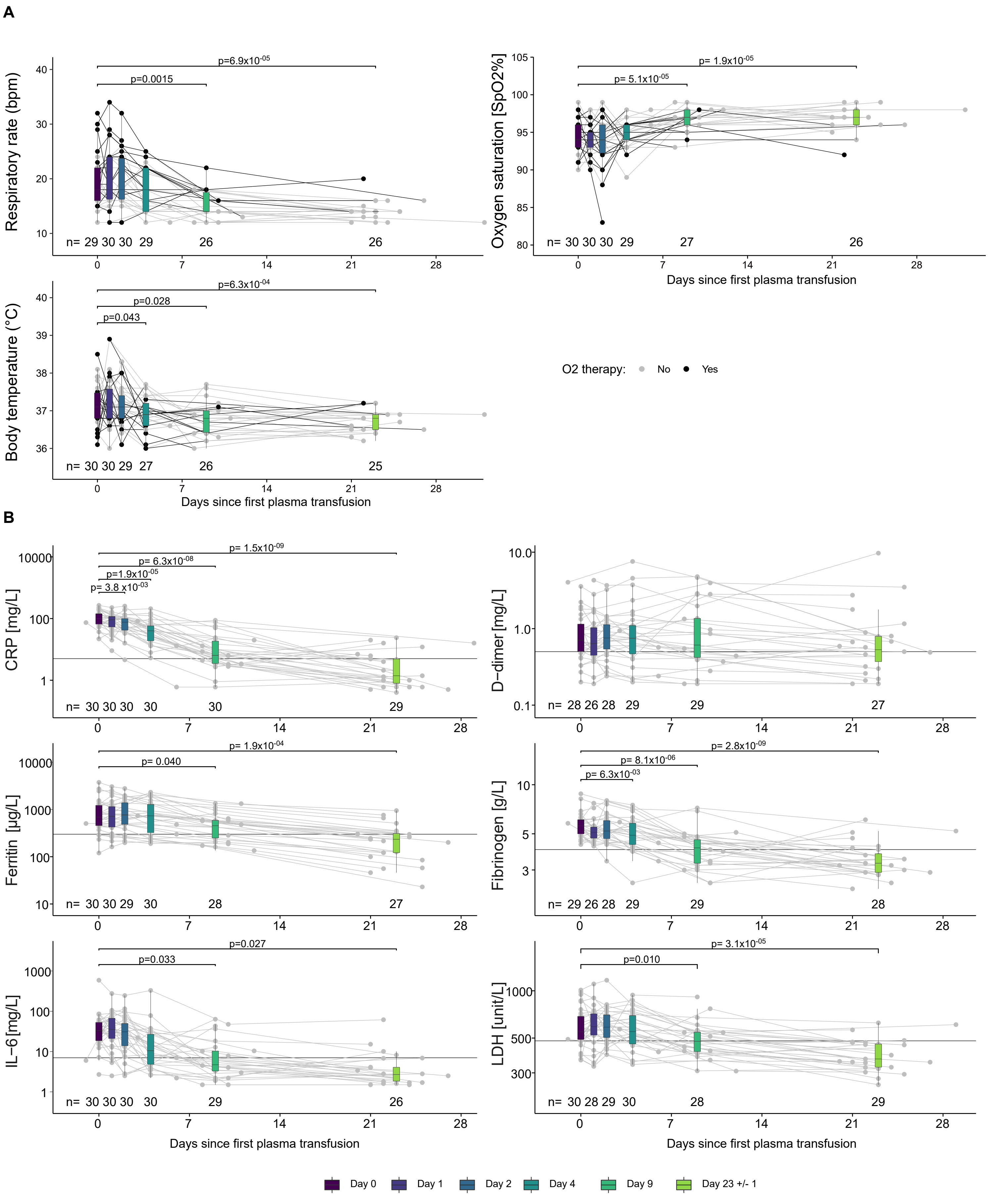
A
Respiratory rate (bpm)
p=6.9x10-05
p=0.0015
n= 29 30 30 29 26 26
Oxygen saturation [SpO2%]
p= 1.9x10-05
p= 5.1x10-05
n= 30 30 30 29 27 26
Days since first plasma transfusion
Body temperature (°C)
p=6.3x10-04
p=0.028
p=0.043
n= 30 30 29 27 26 25
Days since first plasma transfusion
O2 therapy: No Yes
B
CRP [mg/L]
p= 1.5x10-09
p= 6.3x10-08
p=1.9x10-05
p= 3.8 x10-03
n= 30 30 30 30 30 29
D−dimer [mg/L]
n= 28 26 28 29 29 27
Ferritin [µg/L]
p= 1.9x10-04
p= 0.040
n= 30 30 29 30 28 27
Fibrinogen [g/L]
p= 2.8x10-09
p= 8.1x10-06
p= 6.3x10-03
n= 29 26 28 29 29 28
IL−6 [mg/L]
p=0.027
p=0.033
n= 30 30 30 30 29 26
LDH [unit/L]
p= 3.1x10-05
p=0.010
n= 30 28 29 30 28 29
Days since first plasma transfusion
Days since first plasma transfusion
Day 0
Day 1
Day 2
Day 4
Day 9
Day 23 +/- 1

Figure S2

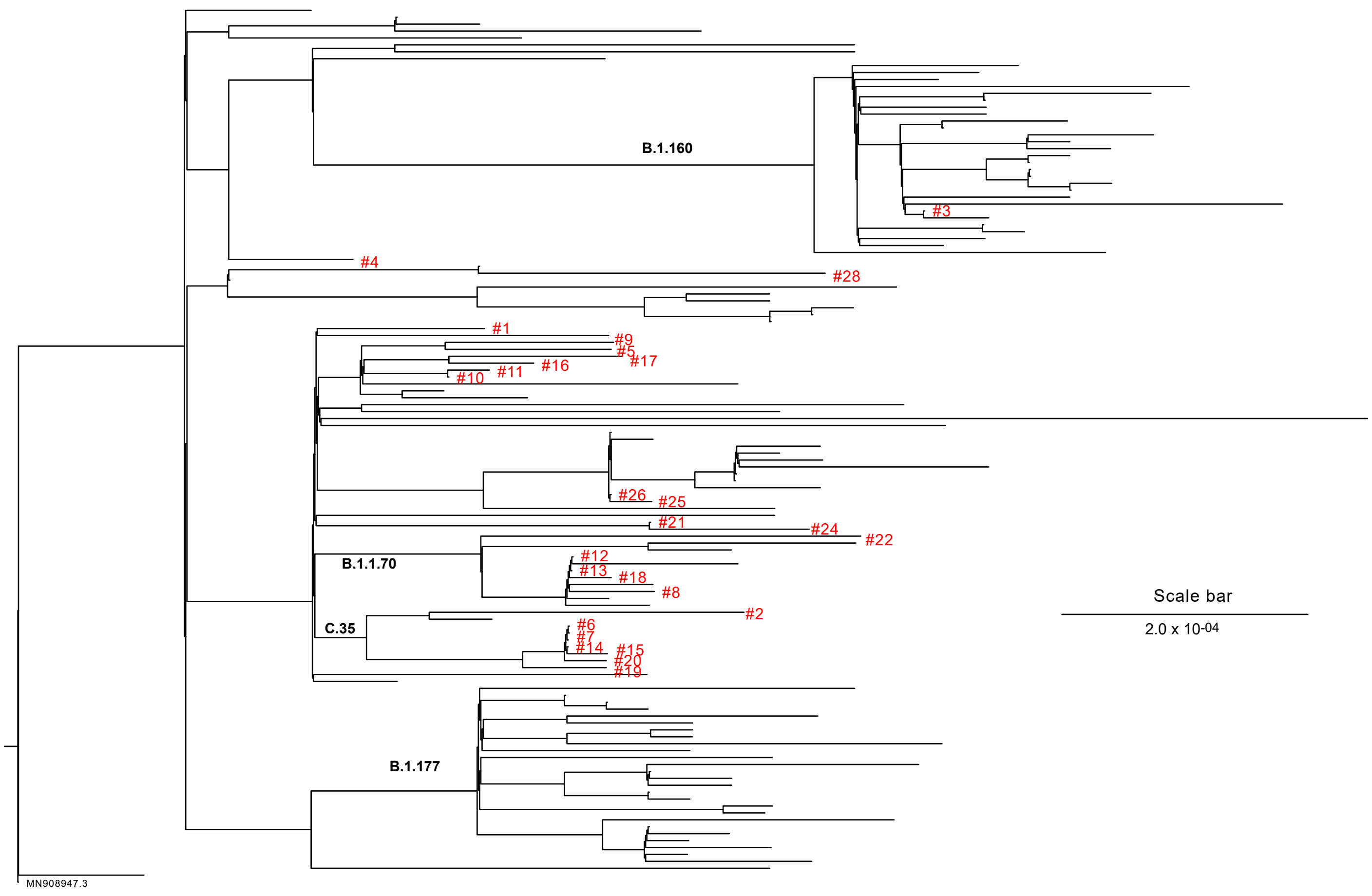
B.1.160
#3
#4
#28
#1
#9
#5
#17
#16
#11
#10
#26
#25
#21
#24
#22
B.1.1.70
#12
#13
#18
#8
C.35
#2
#6
#7
#14
#15
#20
#19
B.1.177
MN908947.3
Scale bar
2.0 x 10-04

Figure S3

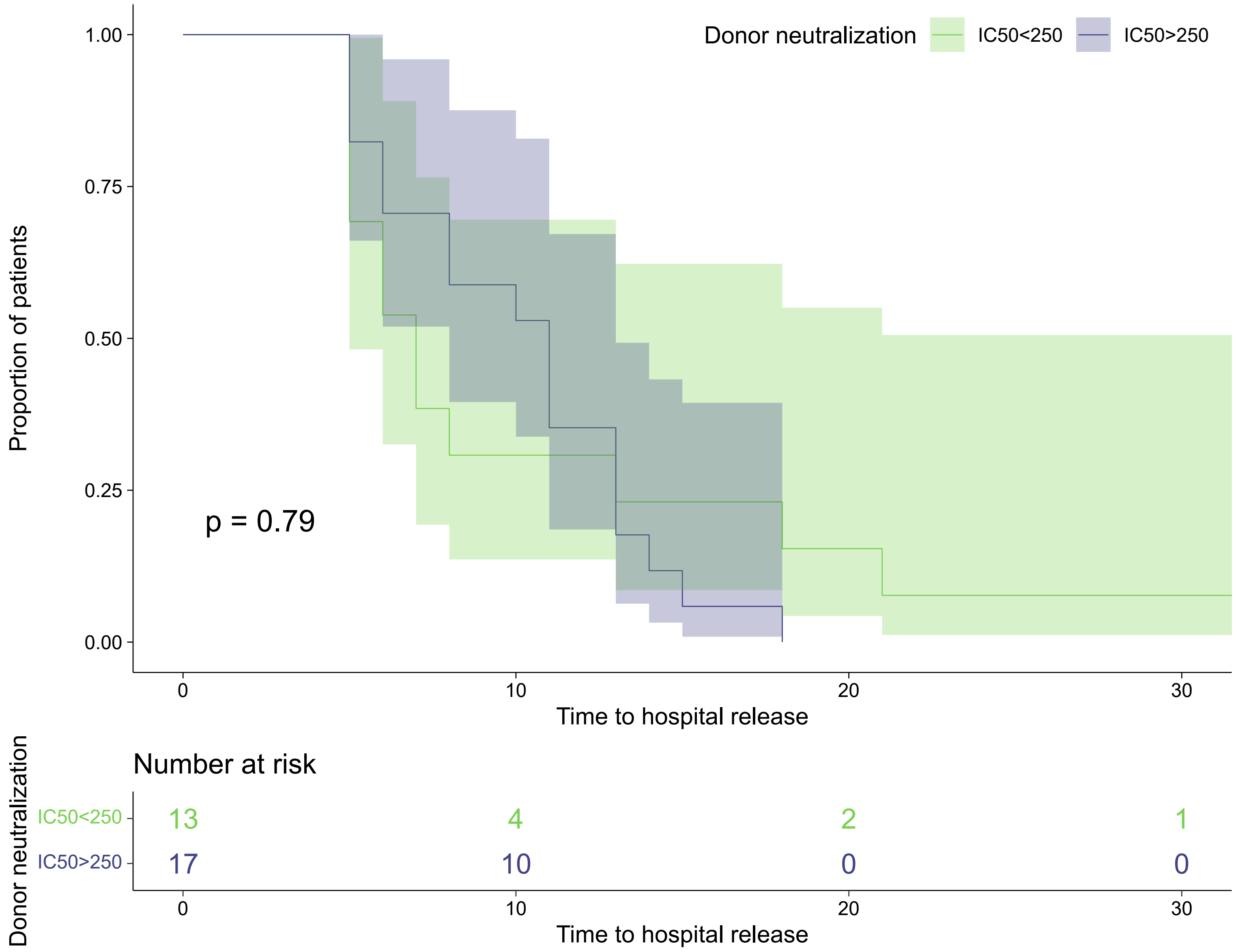
Donor neutralization
IC50<250
IC50>250
Proportion of patients
1.00
0.75
0.50
0.25
0.00
p = 0.79
0
10
20
30
Time to hospital release
Number at risk
Donor neutralization
IC50<250
IC50>250
13
4
2
1
17
10
0
0
0
10
20
30
Time to hospital release

### Figure S4

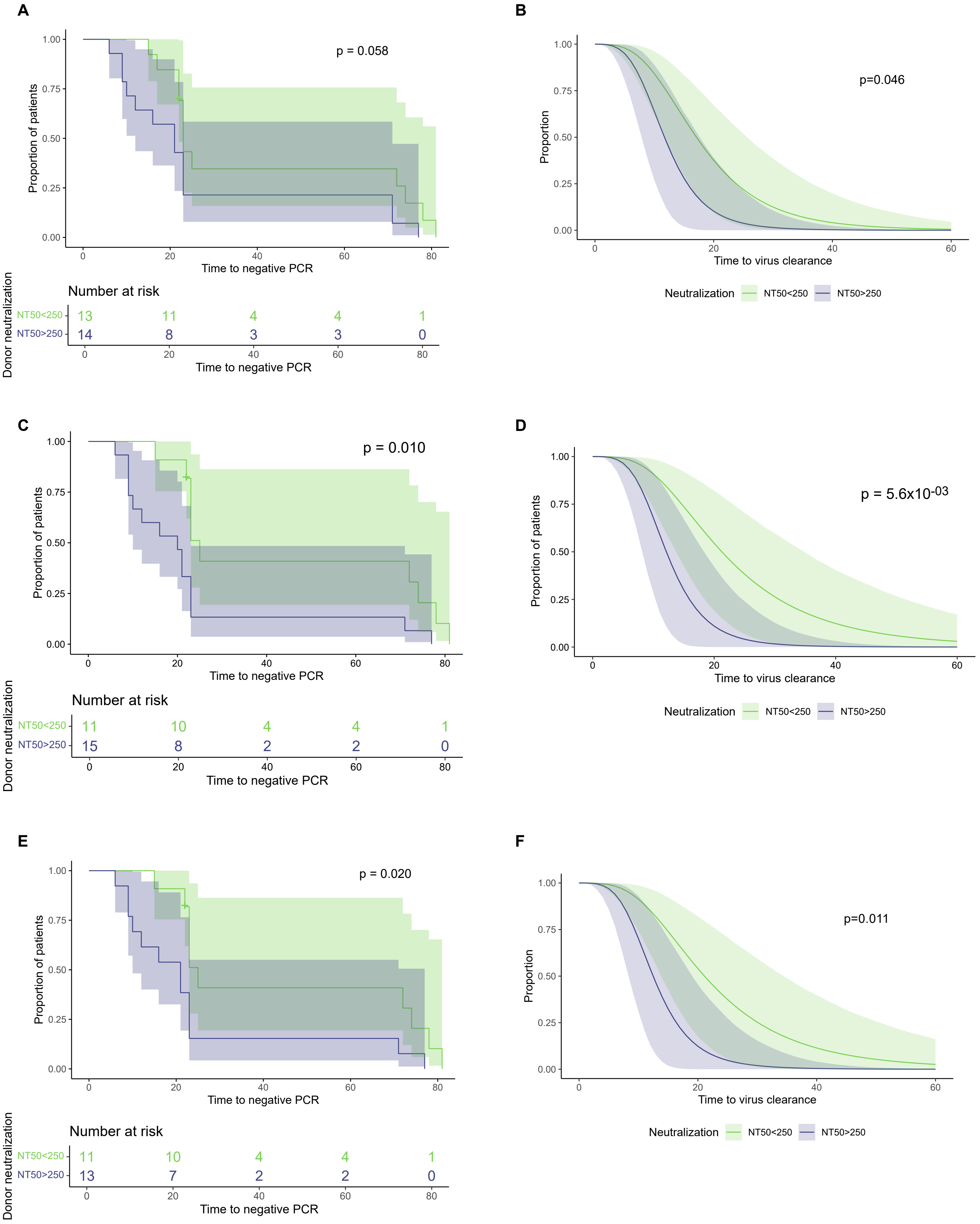

A
p = 0.058
Proportion of patients
1.00
0.75
0.50
0.25
0.00
0
20
40
60
80
Time to negative PCR
Number at risk
Donor neutralization
NT50<250 13 11 4 4 1
NT50>250 14 8 3 3 0
Time to negative PCR
B
p=0.046
Proportion
Time to virus clearance
Neutralization
NT50<250
NT50>250
C
p = 0.010
Proportion of patients
Time to negative PCR
Number at risk
Donor neutralization
NT50<250 11 10 4 4 1
NT50>250 15 8 2 2 0
Time to negative PCR
D
p = 5.6x10-03
Proportion of patients
Time to virus clearance
Neutralization
NT50<250
NT50>250
E
p = 0.020
Proportion of patients
Time to negative PCR
Number at risk
Donor neutralization
NT50<250 11 10 4 4 1
NT50>250 13 7 2 2 0
Time to negative PCR
F
p=0.011
Proportion
Time to virus clearance
Neutralization
NT50<250
NT50>250

### Figure S5

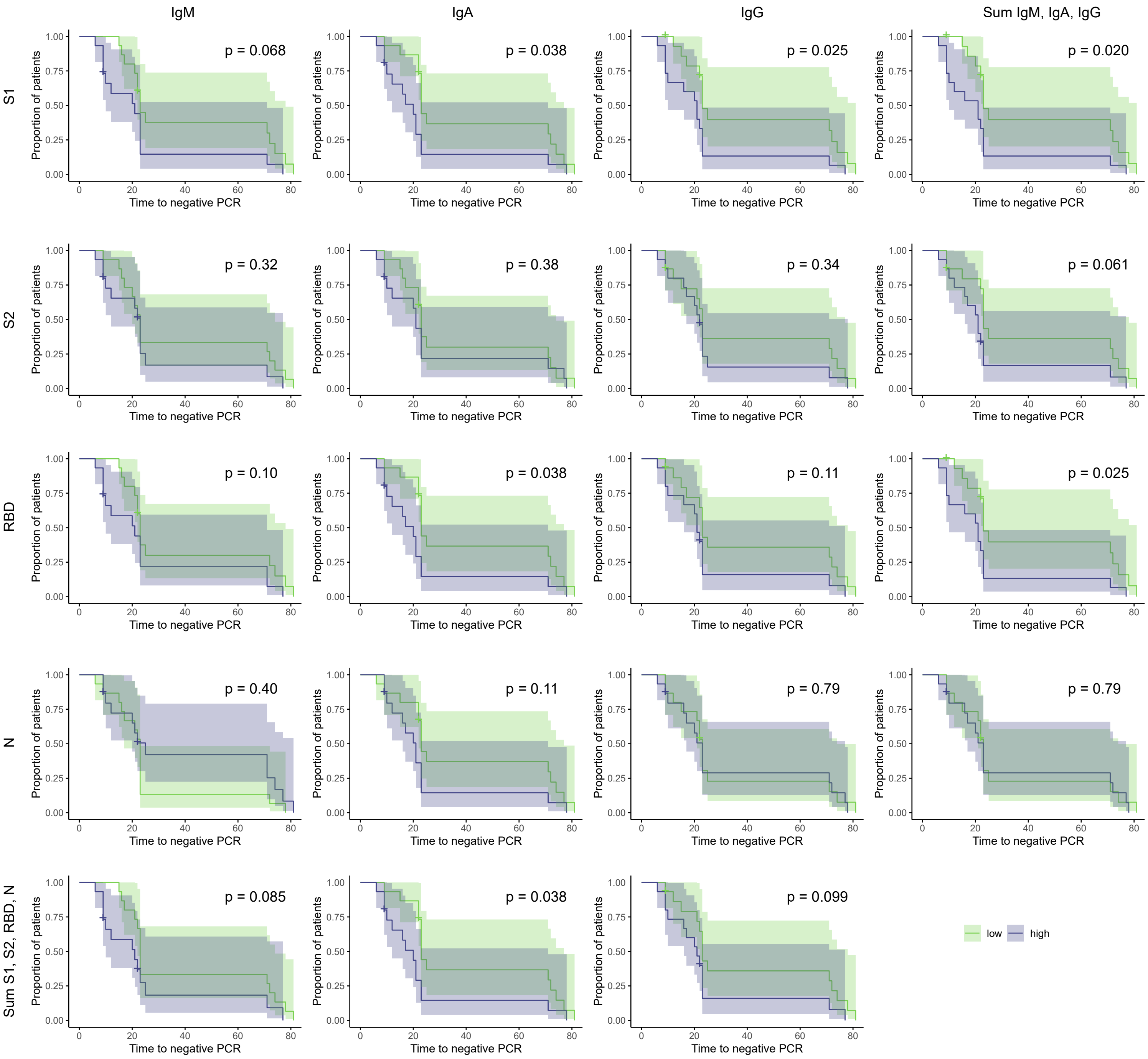
IgM
IgA
IgG
Sum IgM, IgA, IgG
S1
S2
RBD
N
Sum S1, S2, RBD, N
p = 0.068
p = 0.038
p = 0.025
p = 0.020
p = 0.32
p = 0.38
p = 0.34
p = 0.061
p = 0.10
p = 0.038
p = 0.11
p = 0.025
p = 0.40
p = 0.11
p = 0.79
p = 0.79
p = 0.085
p = 0.038
p = 0.099
Proportion of patients
Time to negative PCR
low
high

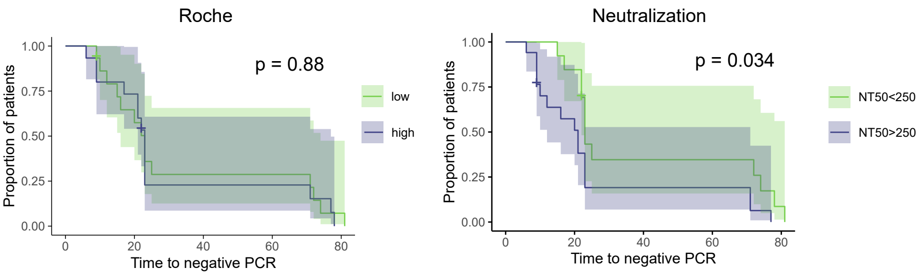
Roche
p = 0.88
low
high
Neutralization
p = 0.034
NT50<250
NT50>250
Proportion of patients
Time to negative PCR

### Figure S6

| Variables | Baseline | Subcategory | HR [95%CI] | p–value |
|---|---|---|---|---|
| Baseline viral load | | + 1 log10(VL) | 0.53 [0.34,0.84] | $6.4x10^{-03}$ |
| | | | 0.46 [0.27,0.78] | $3.6x10^{-03}$ |
| Comorbidity | No comorbidity | At least 1 comorbidity | 0.56 [0.22,1.4] | 0.23 |
| | | | 0.91 [0.33,2.5] | 0.85 |
| Roche S | low | high | 1 [0.42,2.4] | 1 |
| | | | 2.1 [0.75,5.6] | 0.16 |

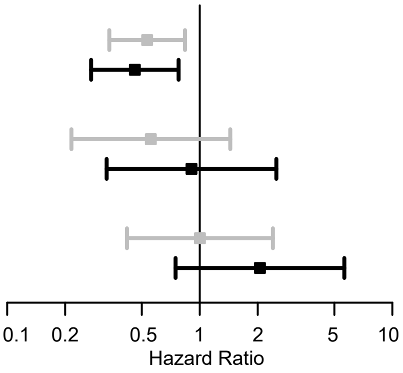


univariable  multivariable

Figure S7

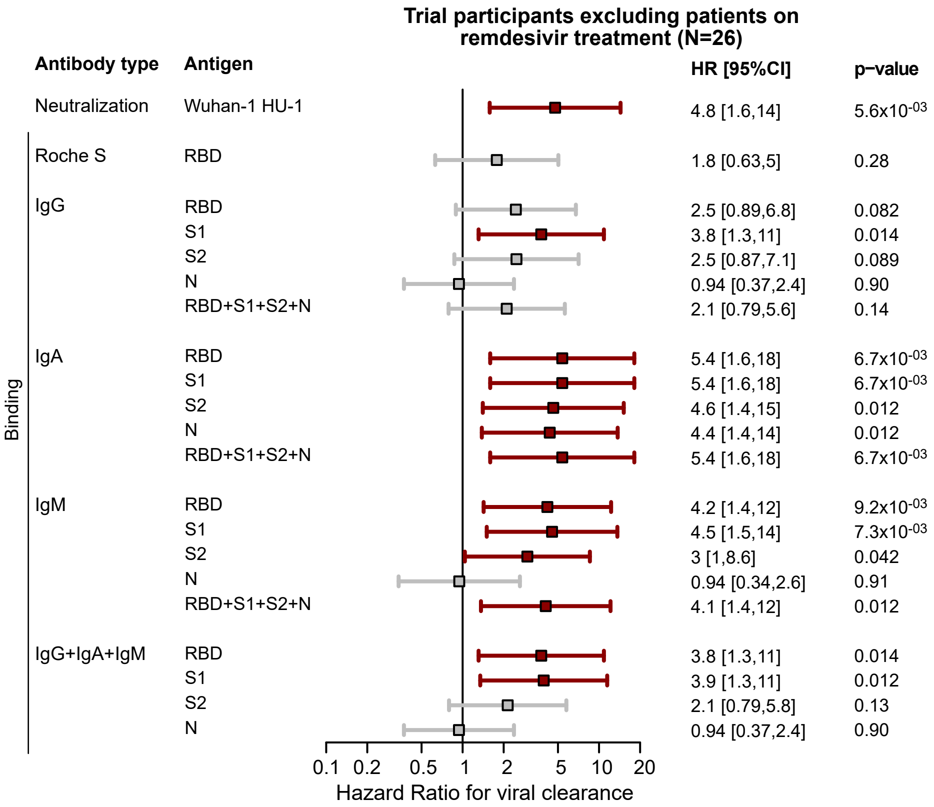
Trial participants excluding patients on remdesivir treatment (N=26)
Antibody type
Antigen
HR [95%CI]
p–value
Neutralization
Wuhan-1 HU-1
4.8 [1.6,14]
5.6x10-03
Binding
Roche S
RBD
1.8 [0.63,5]
0.28
IgG
RBD
2.5 [0.89,6.8]
0.082
S1
3.8 [1.3,11]
0.014
S2
2.5 [0.87,7.1]
0.089
N
0.94 [0.37,2.4]
0.90
RBD+S1+S2+N
2.1 [0.79,5.6]
0.14
IgA
RBD
5.4 [1.6,18]
6.7x10-03
S1
5.4 [1.6,18]
6.7x10-03
S2
4.6 [1.4,15]
0.012
N
4.4 [1.4,14]
0.012
RBD+S1+S2+N
5.4 [1.6,18]
6.7x10-03
IgM
RBD
4.2 [1.4,12]
9.2x10-03
S1
4.5 [1.5,14]
7.3x10-03
S2
3 [1,8.6]
0.042
N
0.94 [0.34,2.6]
0.91
RBD+S1+S2+N
4.1 [1.4,12]
0.012
IgG+IgA+IgM
RBD
3.8 [1.3,11]
0.014
S1
3.9 [1.3,11]
0.012
S2
2.1 [0.79,5.8]
0.13
N
0.94 [0.37,2.4]
0.90
0.1 0.2 0.5 1 2 5 10 20
Hazard Ratio for viral clearance

### Figure S8

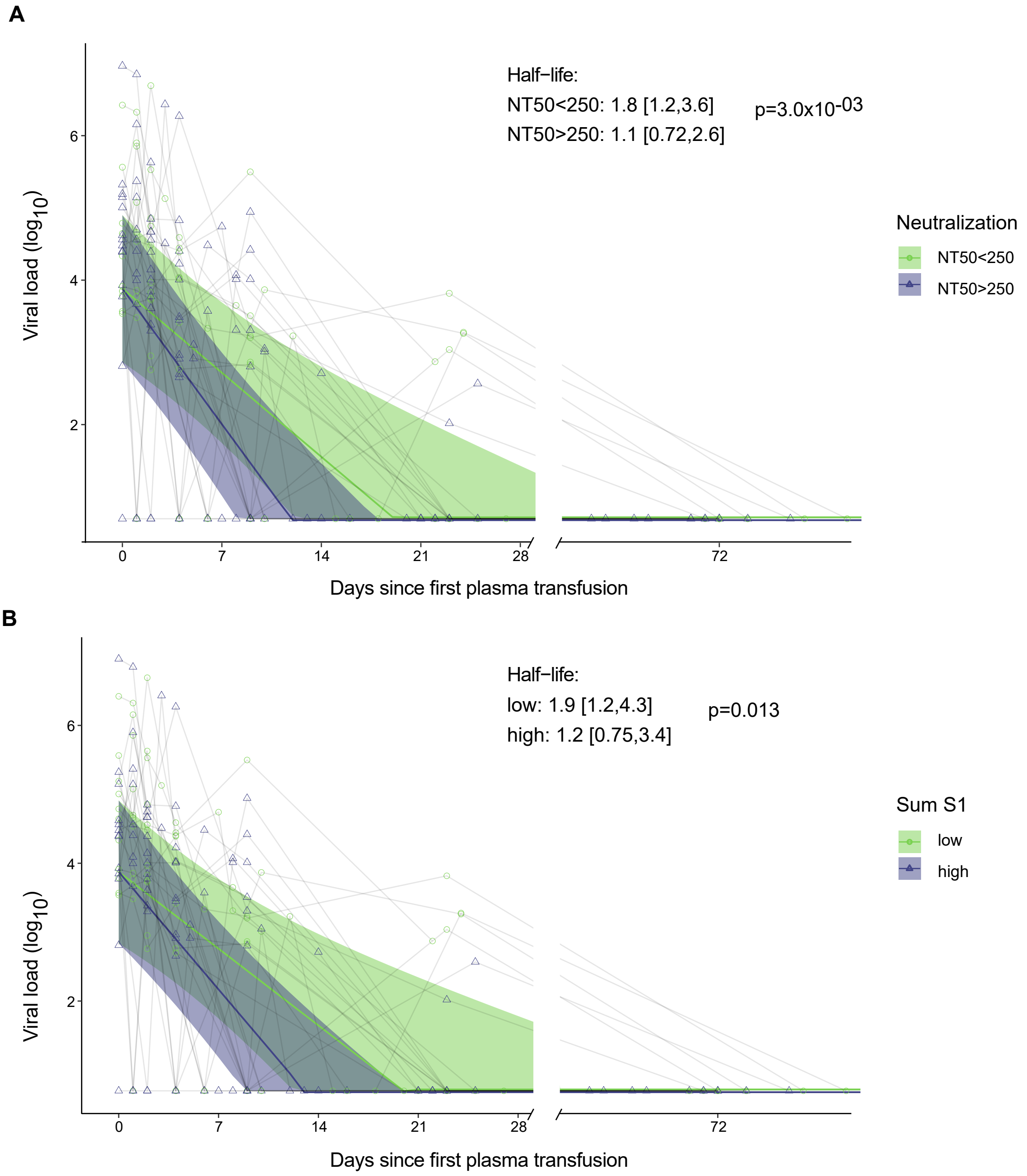
A
Half−life:
NT50<250: 1.8 [1.2,3.6]
NT50>250: 1.1 [0.72,2.6]
p=3.0x10$^{-03}$
Viral load (log$_{10}$)
Days since first plasma transfusion
Neutralization
NT50<250
NT50>250
B
Half−life:
low: 1.9 [1.2,4.3]
high: 1.2 [0.75,3.4]
p=0.013
Viral load (log$_{10}$)
Days since first plasma transfusion
Sum S1
low
high

### Figure S9

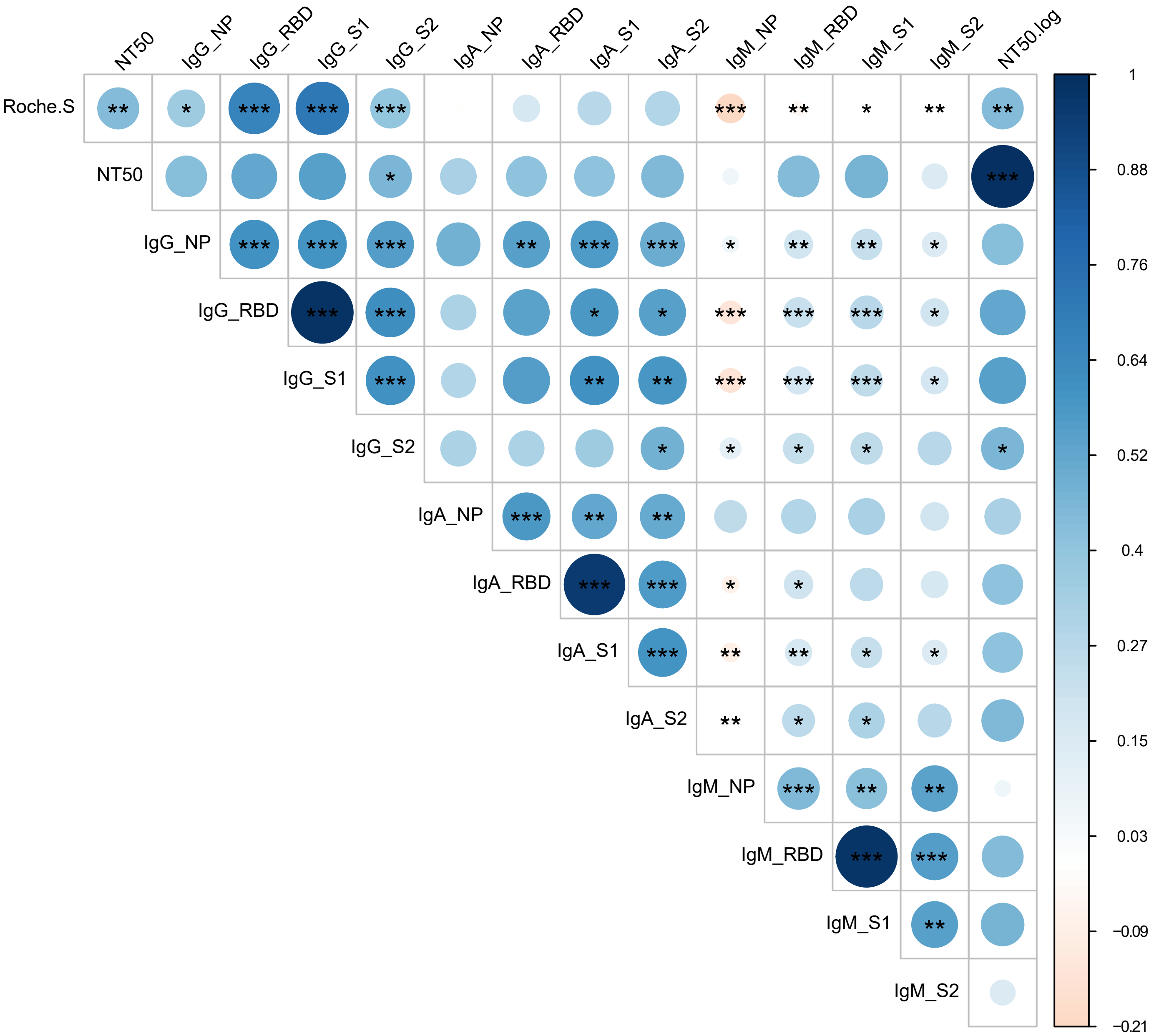
NT50
IgG_NP
IgG_RBD
IgG_S1
IgG_S2
IgA_NP
IgA_RBD
IgA_S1
IgA_S2
IgM_NP
IgM_RBD
IgM_S1
IgM_S2
NT50.log
Roche.S
1
0.88
0.76
0.64
0.52
0.4
0.27
0.15
0.03
−0.09
−0.21

Figure S10

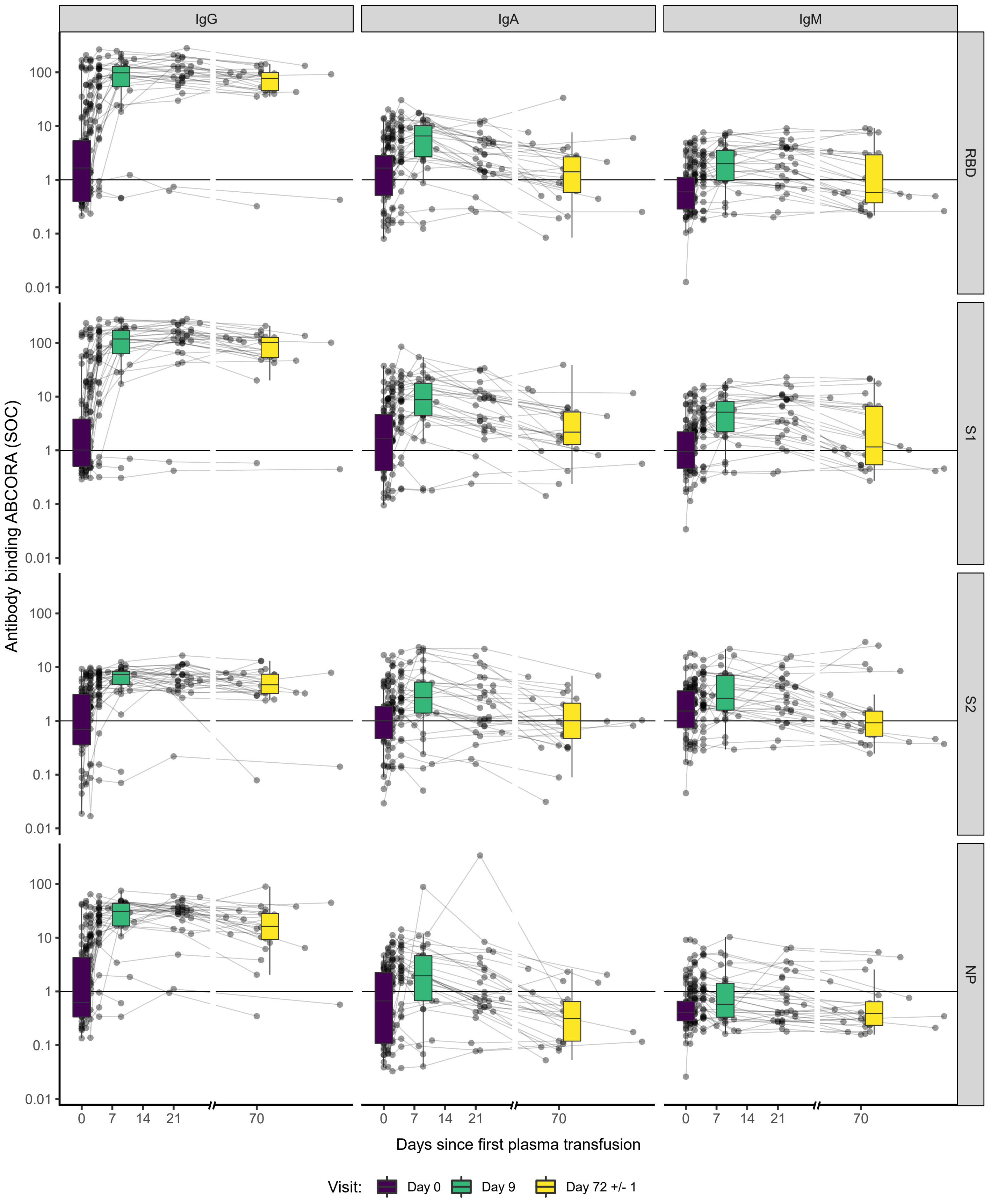
IgG
IgA
IgM
RBD
S1
S2
NP
Antibody binding ABCORA (SOC)
100
10
1
0.1
0.01
0
7
14
21
70
Days since first plasma transfusion
Visit:
Day 0
Day 9
Day 72 +/- 1

### Figure S11

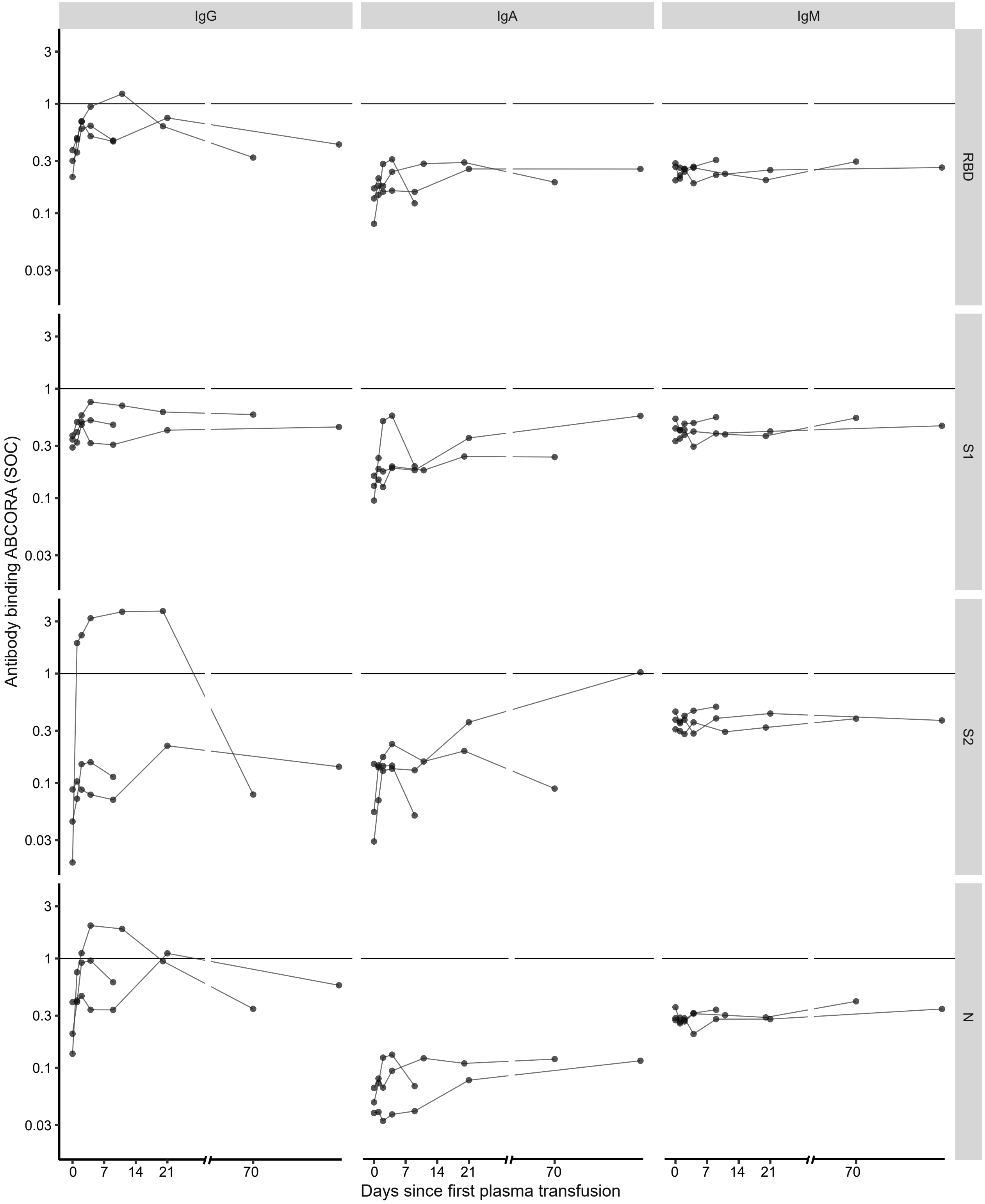
IgG
IgA
IgM
RBD
S1
S2
N
Antibody binding ABCORA (SOC)
3
1
0.3
0.1
0.03
0
7
14
21
70
Days since first plasma transfusion

Figure S12

**A**

| Variables | Baseline | Subcategory | HR [95%CI] | p−value |
|---|---|---|---|---|
| Baseline viral load | | + 1 log10(VL) | 0.5 [0.31,0.8]<br>0.42 [0.23,0.76] | 4.1x10$^{-03}$<br>3.9x10$^{-03}$ |
| Comorbidity | No comorbidity | At least 1 comorbidity | 0.62 [0.21,1.8]<br>0.77 [0.21,2.8] | 0.39<br>0.69 |
| Neutralization (recipients) | NT50=100 | NT50>100 | 0.66 [0.25,1.8]<br>0.42 [0.12,1.5] | 0.41<br>0.18 |
| Neutralization (donors) | NT50<250 | NT50>250 | 2.9 [1.1,7.6]<br>5.2 [1.6,17] | 0.036<br>6.7x10$^{-03}$ |

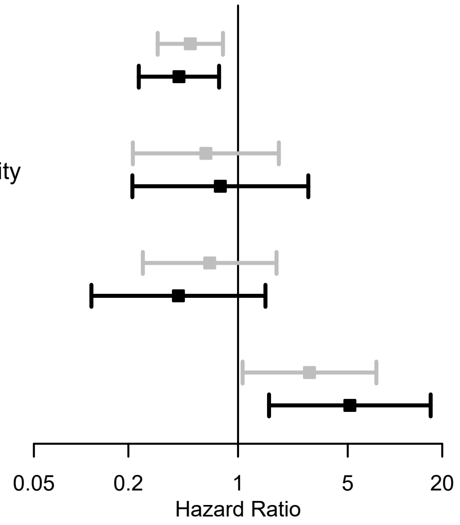


univariable  multivariable

**B**

| Variables | Baseline | Subcategory | HR [95%CI] | p−value |
|---|---|---|---|---|
| Baseline viral load | | + 1 log10(VL) | 0.51 [0.31,0.84]<br>0.5 [0.29,0.85] | 7.6x10$^{-03}$<br>0.011 |
| Comorbidity | No comorbidity | At least 1 comorbidity | 0.44 [0.16,1.2]<br>0.46 [0.14,1.5] | 0.11<br>0.19 |
| Neutralization (recipients) | NT50=100 | NT50>100 | 0.66 [0.25,1.8]<br>0.33 [0.093,1.2] | 0.40<br>0.087 |
| Neutralization (donors) | NT50<250 | NT50>250 | 1.8 [0.69,4.6]<br>3.5 [1.1,11] | 0.23<br>0.034 |

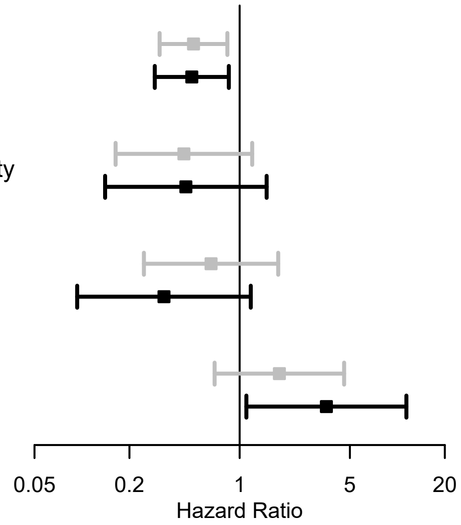


univariable  multivariable

Figure S13

**A**

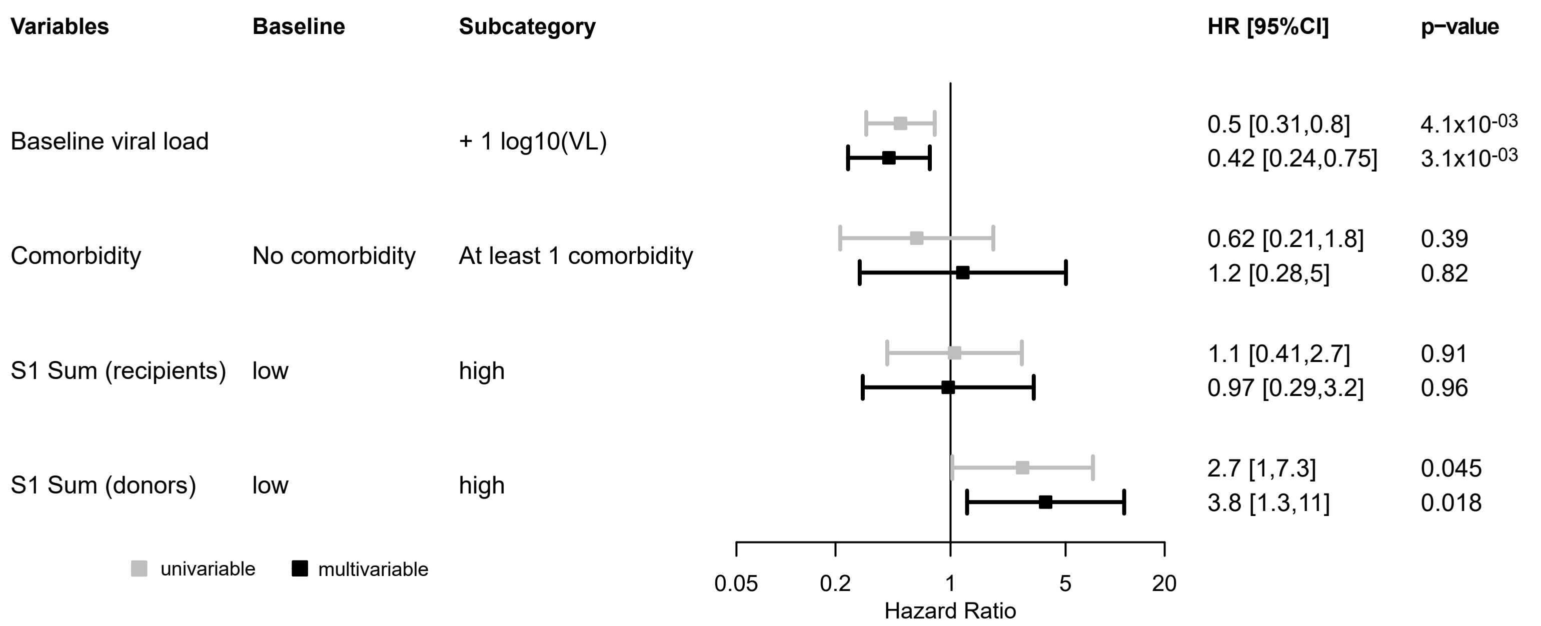


**B**

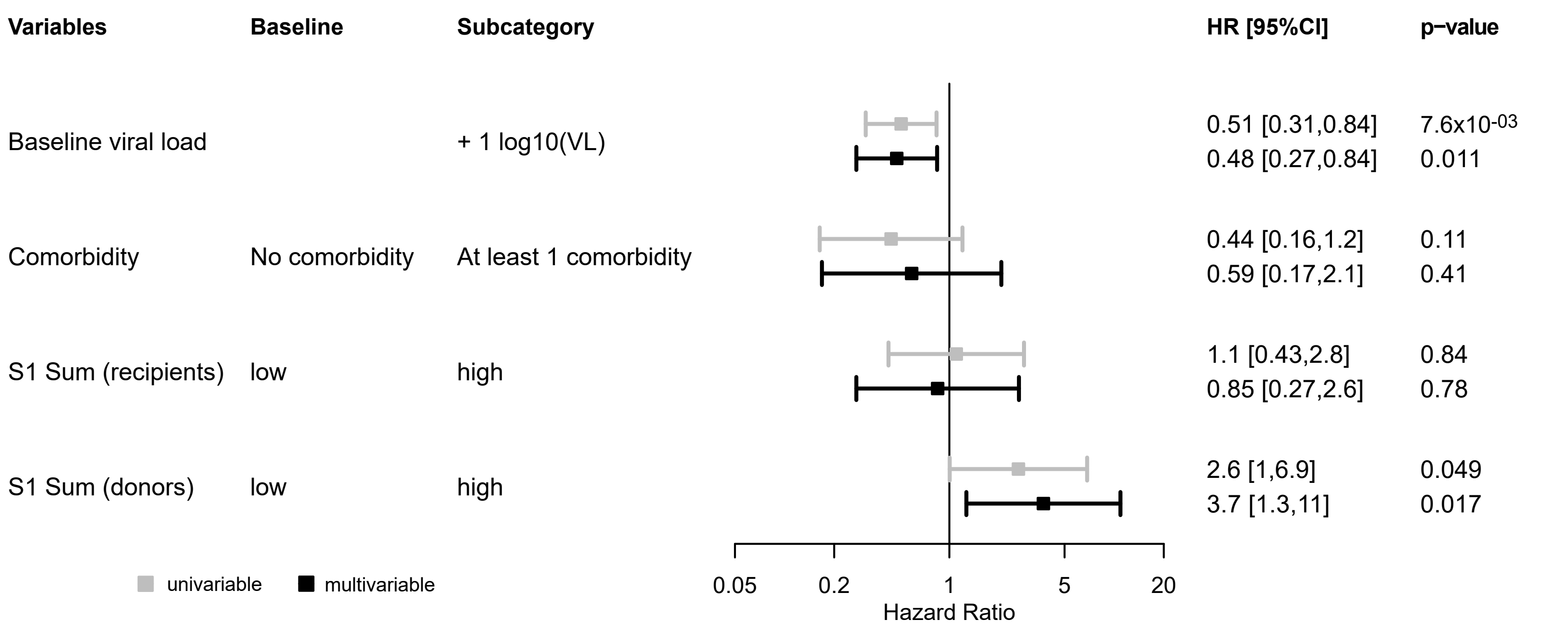
